# A power-based sliding window approach to evaluate the clinical impact of rare genetic variants

**DOI:** 10.1101/2022.07.29.22278171

**Authors:** Elizabeth T. Cirulli, Kelly M. Schiabor Barrett, Alexandre Bolze, Joseph J. Grzymski, William Lee, Nicole L. Washington

## Abstract

Systematic determination of rare and novel variant pathogenicity remains a major challenge, even when there is an established association between a gene and phenotype. Here we present Power Window (PW), a novel sliding window technique that identifies the clinically impactful regions of a gene using population-scale clinico-genomic datasets. By sizing windows based on the number of variant carriers, rather than the number of variants or nucleotides, statistical power is held constant during analysis, enabling the localization of clinical impact as well as the removal of unassociated gene regions. This method can be used to focus on: specific variant types such as loss of function (LoF) or other coding; parts of a gene, such as those expressed in different tissues; or isolating gene regions with opposite directions of effect. Using a training set of 300K exomes from the UKBiobank (UKB), we developed PW-based LoF and coding models for well-established gene-disease associations and tested their accuracy in two additional cohorts (128k exomes from the UKB and 30k exomes from the Healthy Nevada Project (HNP)). The significant PW models retained a mean of 64% of the rare variant carriers in each gene (range 16-98%), with quantitative traits showing a mean effect size improvement of 48% compared to aggregating rare variants across the entire gene, and the odds ratios for binary traits improving by a mean of 2.4-fold. PW showcases that EHR-based statistical analyses can accurately distinguish between novel coding variants that will have high phenotypic penetrance in a population and those that will not, unlocking new potential for population genetic screening.

## Introduction

Statistical analyses of rare genetic variants in large populations present unique challenges. Variants that are only observed in a handful of people, or even one person, lack statistical power to identify whether they are associated with a trait. Gene-based collapsing methods navigate this problem by grouping together similar rare variants, often by predictions of functional consequence, to improve power. However, not all non-synonymous variants in a gene, even when they have the same predicted functional consequence, can be expected to have the same effect on a phenotype, and grouping them together in this way dilutes the overall signal.

In addition to comparisons to cellular and model organism functional assays, many analysis approaches have been developed to identify and prioritize the types of rare variants and gene regions that are most important for an association with a particular phenotype. First, analyses of LoF variants (loss of function: nonsense, frameshifts, and essential splice sites) and coding variants (damaging missense and in-frame indels) are often performed separately, to distinguish their effects. Algorithms like SKAT allow genetic variants in a single gene to have different effect sizes and directions of effect, giving an overall signal for the gene even when not all variants behave the same^1^. Other studies have focused on just analyzing the intolerant regions of the gene or specific gene domains to identify the source of a gene’s signal^2,3^. However, an unbiased statistical analysis method that effectively selects the parts of a gene to include in association studies has not yet been successfully developed and applied to improve the overall statistical associations for rare coding variants.

Here, we present Power Window (PW), a novel technique that leverages paired clinical phenotypes with exome sequences to identify regions of a gene where rare nonsynonymous variants of any type–LoF or coding–are statistically significantly associated with a trait. We use PW to build regional LoF and coding models for well-established gene-disease associations and test these refinements in two additional cohorts. Not only do these models replicate, but many drive dramatic improvements to the effect size or odds ratio, especially for coding variants. PW showcases that even in the absence of family data, prior clinical evidence for that variant, or functional tests, EHR-based statistical analyses alone can determine which type of novel coding variants will have high penetrance in population cohorts. This highly accurate method for prioritizing genetic variants associated with health outcomes unlocks new potential for common disease genomic risk screening.

## Methods

### Genetic data and phenotypes

We utilized the UK Biobank (UKB) plink-formatted population level exome OQFE exome files (field 23155) as well as the imputed genotypes from GWAS genotyping (field 22801-22823). We also utilized 28,423 Healthy Nevada Project (HNP) samples that were sequenced and analyzed at Helix using the Exome+® assay as previously described ^3^. For the UKB cohort (n with exomes=450k), participants range in age as of 2021 from 50 to 87 and are 55% female, while the HNP age range is from 18 to 89+ and is 68% female. The UKB is 83% British European ancestry, with another 10% of other European ancestry and 7% of other ancestries, and the HNP is 77% of general European ancestry, 14% of Hispanic ancestry, and 9% of other ancestries. No filtering was applied to the cohorts based on ancestry.

HNP phenotypes were processed from Epic/Clarity Electronic Health Records (EHR) data as previously described^3^. Quantitative phenotypes were transformed via rank-based inverse normal transformation. UKB data were obtained from the UKB resource. UKB phenotypes were processed using the Neale lab modified version of PHESANT as previously described, which rank-transforms quantitative traits to normally distributed data and divides categorical traits into binary sets^3–5^.

ICD codes were translated to phecodes as previously described^6–9^. For HNP, International Classification of Diseases, Ninth and Tenth Revision ICD codes and associated dates (ICD-9 and ICD-10-cm) were collected from available diagnosis tables (from problem lists, medical histories, admissions data, surgical case data, account data, claims and invoices). For UKB, ICD codes and associated dates (both ICD-9 and ICD-10) were collected from inpatient data (category 2000), cancer register (category 100092) and first occurrences (category 1712) (**Table S1**).

### Genetic analysis

Variant annotation was performed with VEP 99 ^10^. Coding regions were defined according to Gencode version GENCODE 33, and the Ensembl canonical transcript was used to determine variant consequence ^11,12^. Variants were restricted to CDS regions and essential splice sites. Genotype processing was performed in Hail 0.2.54-8526838bf99f^13^.

The collapsing analysis was performed as previously described^6^. Samples were coded as a 1 for each gene if they had a qualifying variant and a 0 otherwise. We defined “qualifying” as coding (missense_variant, inframe_deletion, or inframe_insertion) and not Polyphen or SIFT benign (Polyphen benign is <0.15, SIFT benign is >0.05) or loss of function (LoF) (stop_lost, start_lost, splice_donor_variant, frameshift_variant, splice_acceptor_variant, or stop_gained)^14,15^. We used a MAF cutoff of 0.1%. To pass the MAF filter, the variant must be below that frequency cutoff in all gnomAD populations as well as locally within each population analyzed ^16^. For visualization, UniProt was used to identify gene domains^17^.

We used regenie for genetic analyses^18^. Briefly, this method builds a whole genome regression model using common variants to account for the effects of relatedness and population stratification, and it accounts for situations where there is an extreme case-control imbalance, which can lead to test statistic inflation with other analysis methods. The covariates we included were age, sex, age*sex, age*age, sex*age*age, and bioinformatics pipeline version as appropriate. As previously described, a representative set of 184,445 coding and noncoding LD-pruned, high-quality common variants were identified for building the whole genome regression model^3^.

We analyzed all ancestries together because when collapsing rare (MAF<0.1%) causal variants across a gene and analyzing with a linear mixed model or whole genome regression, signals tend to be consistent whether restricting to one ancestry or analyzing across all ancestries^3^. This method works in this setting because analyses of collapsed rare variants are less influenced by ethnic background than are analyses of the common variants used in a typical GWAS, in large part because causal variants are being grouped together as opposed to tagging variants.

### Training Power Window models

PW uses a sliding window methodology to create windows with identical numbers of rare variant carriers across the gene (**Figure 1A**). To train, we built windows in 300k UKB samples; this otherwise randomly selected set (of the 450k UKB exome release) included all individuals who were first or second-degree relatives or twins, so that the subsequent testing set would only include unrelated individuals. When building the windows, we required them to each contain 20 qualifying variant carriers out of these 300k individuals (see **Figure S1** for stats about the windows). PW analyses were run for 65 established gene/disease relationships (see Results). Specifically, we built the windows separately for LoF and coding models, and we only built windows for gene/model combinations that had at least 25 qualifying rare variant carriers in the training set (this excluded 7 genes (*CDKN2A, FCGRT, GCK, GFI1B, GP9, SF3B1*, and *SLC4A1*) from the LoF model and 0 genes from the coding model). We then ran a regression analysis of the relevant phenotype in regenie on this training set for each window generated for each gene.

**Figure 1.**
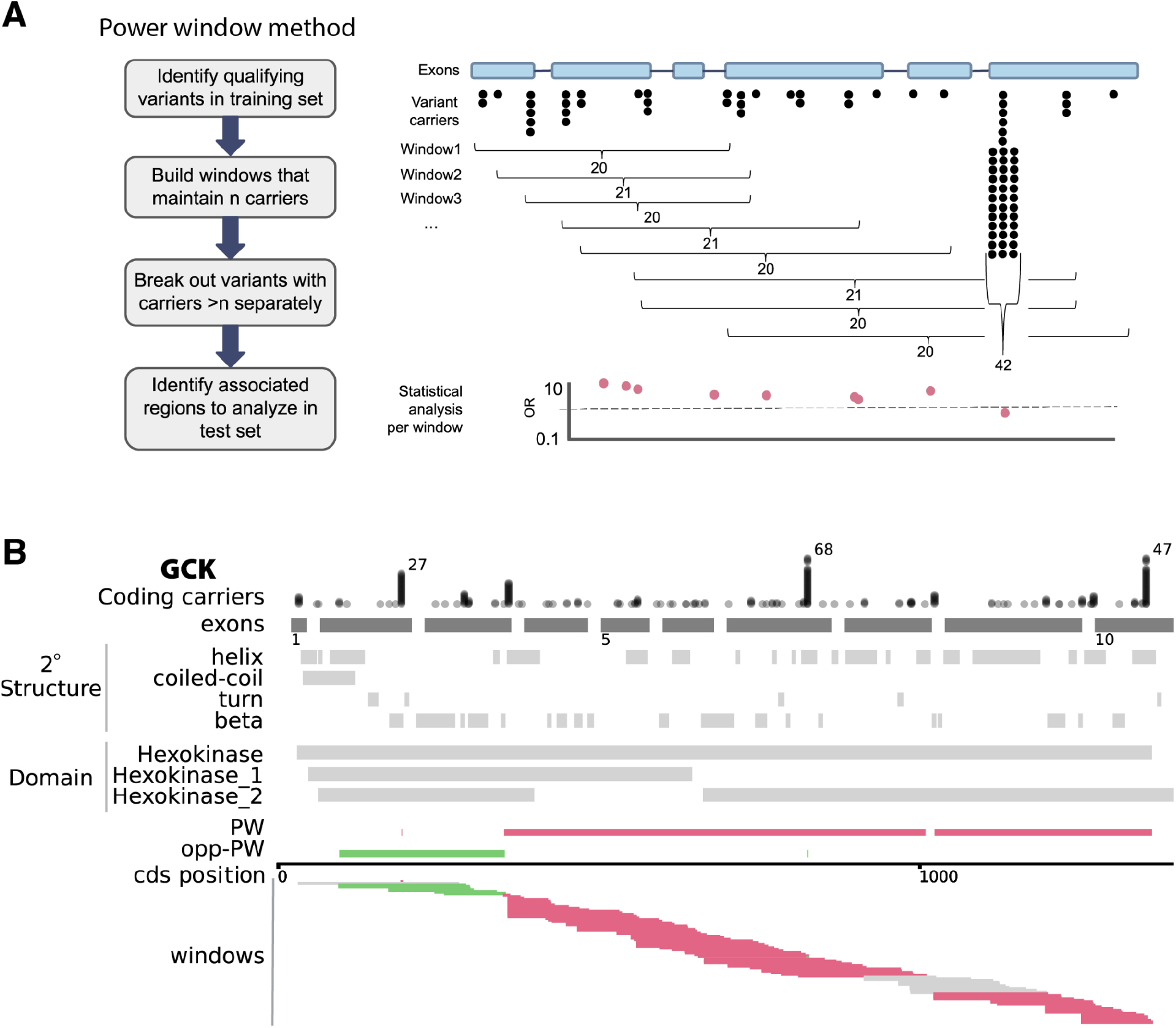
Power Window methodology and as applied to *GCK* in UKB training data. A) Diagram of the methodology. B) Power Window (PW) applied to *GCK* and glucose levels in the UKB300K discovery cohort. Tracks are drawn against the *GCK* canonical coding transcript ENST00000403799.8. Coding position is shown at the bottom scale. Coding carriers: each dot represents an individual carrier and are stacked for each carrier at a given position. For brevity, carriers are trimmed to 35 and total number of carriers is indicated when total carriers>40. *GCK* exons: exons (dark grey to scale; introns not to scale). Exon number indicated below exon track. Secondary structure and major structural domains are shown according to UniProt. PW: bedtools merge of all significant PW_coding_ windows with a positive direction of effect (beta>0.5; pink), as indicated in the ‘windows’ track below. opp-PW: merger of all significant windows with a negative direction of effect (beta<-0.5; green). Windows: each window that was generated through applying the PW algorithm is shown, with a window size of 20 carriers per window. Significant association with glucose levels is indicated when beta<-0.5 or beta>0.5 (99% confidence that the true beta is not 0 in a sample of 20 individuals with a normalized phenotype). Windows are shown in pink if significant under the positive model and green if significant under the negative model, or grey if not significant (beta between -.5 and 0.5).

We next decided which windows to retain in our final model for each gene using a confidence interval approach. For a sample size of 20 carriers measured for a rank-based inverse normal transformed phenotype with a true mean of 0, then 99% of the time, the observed mean in the sample will be between -0.5 and 0.5. An effect size >|0.5| was therefore chosen as the cutoff for a window to be considered associated with the quantitative trait. For binary traits, the cutoff was an odds ratio higher than the maximum expected 99% of the time if the true odds ratio were 1 in a sample of 20, given binomial probability and the case frequency for that phenotype. For example, if the true odds ratio (OR) were 1, then in a sample of 20 carriers, 99% of the time a binary trait occurring in 1 out of 100 people would be expected to have fewer than 3 case carriers and thus an OR<17.5 when compared to the 299,980 non-carriers. If the binary trait occurred in 1 out of 20 people, then the OR cutoff would be 6.3 (5+ case carriers). The odds ratio cutoff was thus different for each binary trait and tailored to its frequency, including tailored to sex-specific analyses when necessary, such as for breast cancer.

The final PW model for each gene separated the gene into regions that had statistical evidence for an association with the trait (PW) and those that did not (non-PW). For quantitative traits, sometimes two directions of effect were observed within different regions of the same gene, in which case an additional model was produced for the regions that had the opposite direction of effect from the main signal for the gene (opp-PW). Models were built separately for coding (PW_coding_) and LoF (PW_LoF_) annotations.

### Testing Power Window in independent samples

We next tested the PW models, non-PW models, and whole gene models in an independent set of 128k unrelated UKB samples. Individuals were analyzed separately according to whether they had variants within the PW regions, within the non-PW regions, or, for quantitative traits, within the opp-PW regions. We compared the ORs and betas in this independent testing cohort between the whole-gene model, the PW model, and the non-PW model, where little signal is expected to remain if the method is appropriately picking out the associated portions of the gene. We considered PW models to be significant in the test set if 1) the 95%CIs of the PW and non-PW (or PW-opp) in the test set did not overlap each other (quantitative and binary traits); 2) the 95%CIs of the PW model did not overlap -0.5 to 0.5 (quantitative traits); 3) the OR was higher than the OR threshold used to select windows for the model in the training set (binary traits); and 4) there was more than 1 case carrier in the PW model for the test set (only 1 model failed this criterion: the negative control model for *IQGAP2* PW_LoF_).

For the significant PW models, we additionally performed a replication study in an independent set of 28k samples from the HNP. For binary traits, we only tested the significant models in HNP if there were at least 5 case carriers expected in the HNP cohort (determined by UKB case carrier frequency * HNP n cases). Phenotypes were available to check and had enough sample size in this secondary cohort for 49 of the models.

## Results

### A statistical power-based sliding window method to localize rare variant association signals within a gene

The basic concept of a sliding window analysis is to group variants located near each other into one unit and analyze them together to improve power, much like a gene-based collapsing analysis but at a more localized scale. Rather than size our sliding window by the number of variants or bases covered, our sliding window moves to maintain roughly the same number of people within the cohort with a rare qualifying variant, and thus the statistical power, within each window (**Figure 1A**). When a single variant is well powered on its own, it is removed out into its own separate analysis, and the window slides past it, continuing to group surrounding variants as appropriate. We call this technique the Power Window (PW) method.

### Parameters and functions

The carrier count used to define windows can be adjusted to fit different scenarios. There is a balance to strike, rooted in the currently available cohort sample size, between being able to home in on a specific region (small window size) and having adequate power to observe statistical associations (large window size) (**Figure S2**). We use a window size of 20 qualifying variant carriers in a training set of 300k UKB exomes because this powers us to identify associations with quantitative traits with an effect size >|0.5| for a normalized trait (see Methods). This breaks each gene into a mean of 304 coding windows and 66 LoF windows (**Figure S1**). With the same number of qualifying variant carriers within each window, the statistical power for discovery is the same for each window as our analysis slides across the gene. Bases that fall within the span of an associated window are assigned the same value in new datasets as that window has in the training set. Thus, new mutations of the same predicted impact (LoF, missense, etc.) that do not occur in the training dataset can still be assigned a value based on their location in the new dataset. This also means that regions with no variation in the training set are assigned a value based on the values of carriers in the surrounding regions, which defines whether or not they are within the boundaries of an associated window (for example, the third exon in **Figure 1A**). The ability of this method to home in specific regions of the gene increasingly improves as sample sizes grow.

### Power Window models refine gene-disease association signals

We evaluate the PW method using 65 genes with which we previously demonstrated rare variants to be significantly associated with well-documented phenotypes in the UKB using a gene-based collapsing analysis incorporating LoF or coding variant models^3,6^. We chose these associations because they have strong, well-established effects that can be identified in smaller cohorts than the sample used here. There are 37 quantitative traits and 28 binary traits included in this set. For each gene, we build and analyze both coding and LoF windows, irrespective of the original whole-gene association model, for the relevant phenotype in a training set of 300k UKB exomes and build a final model that separates the gene into PW regions and non-PW regions (see Methods).

We first examine if there are differences in how a PW analysis of coding variants (PW_coding_) or LoF variants (PW_LoF_) separates regions within a gene (**Figure 2**). We anticipate that PW_LoF_ models will usually implicate the entire gene instead of specific regions, as generally a LoF variant anywhere in the gene is likely to have a similar effect (with tissue-specific splicing causing some exceptions, see ^19^). Overall for the 65 genes, we find that applying PW_coding_ retains a mean of 37% of the carriers in each gene (range 2-98%), while PW_LoF_ retains 72% (range 0-100%) of carriers. In terms of the percent of the coding sequence (CDS) retained in the PW model in each gene, this is 44% (range <1-98%) for PW_coding_ and 80% (range 0-100%) for PW_LoF_ (**Figure S3**; the 0% is for *JAK2* with myeloproliferative disease, as this association is known to be entirely due to a single missense somatic mutation (rs77375493)^3,20^. While a small percentage of the gene being retained indicates that the association signal is very specific, PW also identifies the gene-phenotype associations where a rare variant anywhere in the gene appears to have a similar effect, which occurs frequently for PW_LoF_ but also for some PW_coding_ models. We find that 52% of the PW_LoF_ models retain >90% of the CDS of the gene, compared to 11% for PW_coding_.

**Figure 2.**
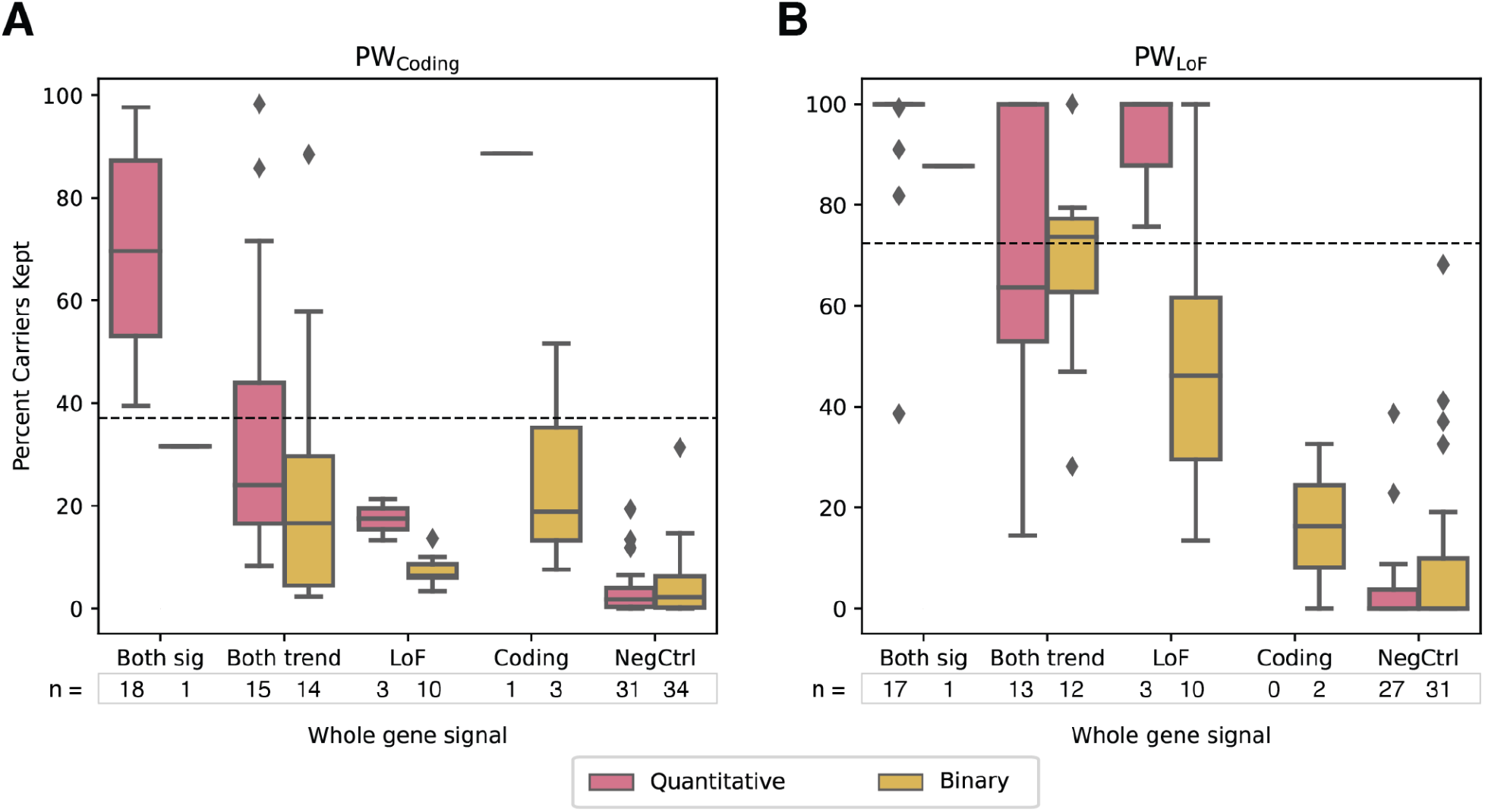
Percent carriers kept in Power Window models. For each gene (n=65), the percent of the rare variant carriers from the whole gene model who were retained by the PW model was evaluated for (A) PW_Coding_ and (B) PW_LoF_. Within each model, the phenotypes were grouped into quantitative (pink) or binary (yellow) traits, and the genes were grouped based on the original genome-wide association as follows: **both sig**: original associations were genome-wide significant for both a coding and LoF models (p<5e-8); **both trend**: genome-wide significance was shown for either coding or LoF, the other model showed at least a nominal association (p<0.05); **LoF**: LoF only and not coding; **coding**: coding only and not LoF; **NegCtrl**: The same genes and phenotypes, but randomly re-assigned to pairs where there is no signal for association with the phenotype at the gene level. For all non-negative control genes, at least one window was included in the final Power Window (PW) model, with the exception of *JAK2* with myeloproliferative disease for the LoF model, which had a whole-gene association signal only for the coding model. For negative control genes, 26% / 24% of the PW_Coding_ / PW_LoF_ models had 0 windows included in the final PW model. Axis label indicates the number of genes tested within each group; note that models were only built if there were at least 25 carriers in the training group, which disqualified 7 genes from having PW_LoF_ models. Dotted horizontal line indicates the mean percent carriers kept (not including NegCtrl models). The percent CDS kept can be found in (**Figure S3**).

To better understand the ways in which PW is able to refine the signal for different gene-phenotype relationships, we group the same 65 genes based on the original model in which the significant association was observed for the whole gene (LoF only, coding only, or both) to see if there are differences in the resulting PW models. We observe that the PW_LoF_ model retains most of its carriers, and thus CDS, in the analysis as long as the original association was not only found with a Coding model (**Figure 2B**). In contrast, for PW_coding_, the portion of the gene that is kept depends heavily on the type of association seen at the whole-gene level (**Figure 2B**): if the whole-gene association signal is only significant for a LoF model (n=13), then a mean of 10% (range 3-21%) of the carriers are retained by the PW_coding_ model, translating to a mean of 18% (range 2-47%) of the CDS. When the whole-gene association can be identified using either a coding model or both coding and LoF models (n=52), then a mean of 44% (range 2-98%) of the carriers are retained, corresponding to a mean of 50% (range 1-98%) of the CDS (**Figure 2A**).

As a negative phenotypic control, we analyze the same genes with the phenotypes swapped so that the models are built with phenotypes that are not associated with the gene. Using the same thresholds as for the main analysis, a mean of 3.8% (range 0-31%) of the variant carriers are retained for PW_coding_ models and 6.1% (0-61%) for PW_LoF_ (**Figure 2**). The percentage of models that include <5% of the gene are 75% / 76% for PW_Coding_ / PW_LoF_, compared to 9% / 2% for the PW models built for phenotypes that show associations at the gene level. The overall distribution of percent carriers retained in the PW models is significantly different between the models built for the negative controls and the main phenotype / gene combination, showing that the PW signal for each gene is indeed specific to the established association (Kruskal-Wallis H-test p=1.5e-33).

### Power Window models dramatically improve odds ratios and effect sizes in new datasets

To confirm the refinements established in the training set of 300k UKB exomes, we evaluate the predictive power of the PW models in an independent testing set of 128k UKB exomes. For each gene, we identify test set individuals with qualifying variants in the regions included in the PW models (PW) or excluded from them (non-PW). This analysis includes a mean of 118 / 27 new Coding / LoF variants per gene, adding to the mean of 542 / 108 Coding / LoF variants already included from the training set (for *TTN*, which we exclude from the summary stats due to its size, the values are 2607 / 308 new and 11958 / 1077 old Coding / LoF variants). We analyze the relevant phenotypes in these groups compared to non-carriers and compare the resulting ORs (binary traits) and effect sizes (betas; quantitative traits) (**Figures 3, S5, S6**). We then identify models that show statistical support for PW as follows.

**Figure 3.**
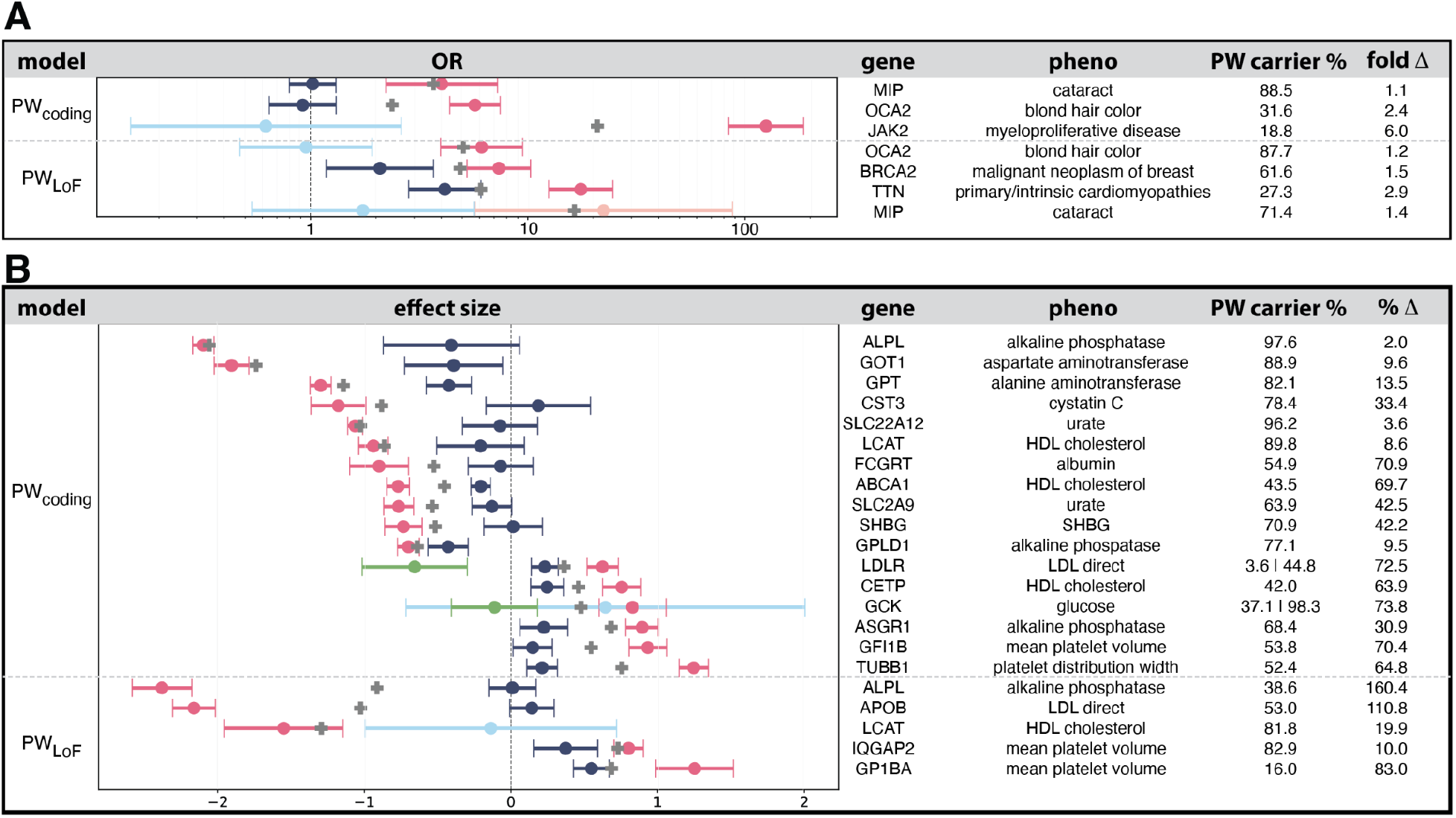
Performance of significant Power Window models in the 128K UKB test set for (A) binary and (B) quantitative traits. Odds ratio (for binary traits in A) and Effect size (for quantitative traits in B) values for gene regions are defined by inclusion in a PW model (PW, pink dot) or exclusion from a PW model (non-PW, blue dot) are plotted against the score for the whole gene (grey cross), together with 95% confidence intervals. Each row is an independently tested gene-phenotype association. Percent of rare variant carriers in the gene that were included in the PW model are indicated. When there is a significant opposite direction of effect within a gene, these are isolated to their own group (opp-PW, green dot) and independently tested, and percent shown is for negative effect / positive effect models. Models are only shown here if the confidence intervals for the PW and non-PW (or opp-PW, meaning there is an opposite direction of effect for some regions of the gene) do not overlap and our criteria for significance were met (see Methods). Remaining models tested, but failing these criteria, are shown in Figures S5 and S6.

For binary traits with PW_coding_, we find that 23 out of 28 gene-disease associations show a higher OR in the PW compared to whole gene model (mean fold improvement=3.7, range=0.14-23.8), and 10 (36%) of these differences are pronounced, with 95%CIs that do not overlap between the PW and non-PW portions of the gene (**Figure 3A** top panel; **S5A**). For example, the association between coding variants in *OCA2* and blonde hair color has an OR of 2.4 (95%CI 2.0-2.8) for the whole gene, but PW_coding_ splits this into 31.6% of carriers with an OR of 5.7 (95%CI 4.4-7.5) in the test set, while the 68.4% that are non-PW carriers have an OR of 0.92 (95%CI 0.6-1.3) (**Table S1**). The regions implicated in this model are all in the citrate transporter domain of this gene, which can be hypothesized to impact its ability to maintain melanosome pH (**Table S2, Figure S4**) ^21^.

For quantitative traits with PW_coding_, the resulting models are even more reliable for the test set: we find that 36 out of 37 genes show a more extreme effect size in the PW compared to whole gene model (mean percent improvement=127%, range=-15 to 836%), and 26 (70%) of these differences have no 95%CI overlap between the PW and non-PW models. For example, the association between coding variants in *GFI1B* and normalized mean platelet volume has an effect size of 0.55 (95%CI 0.46-0.64) for the whole gene, but PW_coding_ splits this into 50.8% of carriers with an effect size of 0.93 (95% CI 0.80-1.06) in the test set, while the 49.2% that are non-PW carriers have an effect size of 0.15 (95% CI 0.01-0.28) (**Table S1**). The associated regions overlap zinc finger domains, which have previously been demonstrated to be involved with this trait (**Figure S4**)^3,22,23^. In addition to having non-overlapping 95% CIs, 17 (49%) of the PW_coding_ models have effect sizes even more extreme than the criteria used to select the windows in the training set (**Figure 3B** top panel; **S6A**; see Methods for details). This was also true for three (11%) of the models for binary traits. We classify these models (n=29; **Figure 3**) as the significant PW models.

Compared to the original whole-gene model, the level of improvement from PW is often quite dramatic: for example, 50% of the significant PW_coding_ models for quantitative traits show a more than 40% improvement in the normalized effect size. Overall, the mean fold improvement for significant PW_coding_ models is 3.2 for ORs (range 1.1-6.0; binary traits) and 40% for percent change in effect size (range 2-74%; quantitative traits) (**Figure 4A** and **4B**, respectively). Additionally, dramatic improvement is observed for some non-significant models as well, but a lower frequency of variant carriers for these models results in lower power to assess significance. For example, the PW_coding_ model for *HBB* with mean corpuscular volume shows 836% improvement over the whole gene model, but as there are only 11 carriers in the PW regions, the confidence intervals overlap those for the non-PW carriers. Further studies with more carriers should clarify the utility of these models.

**Figure 4.**
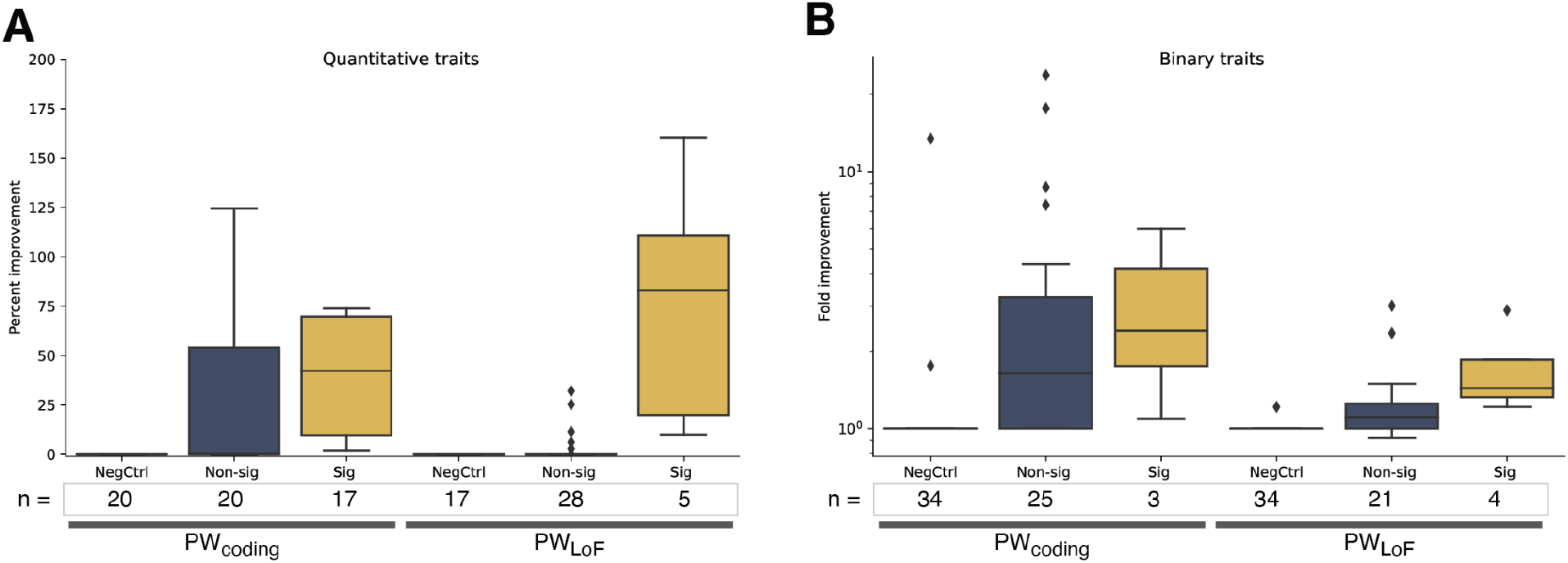
PW-based improvement for normalized traits measured by (A) percent change in effect size or (B) fold improvement in OR. The results are grouped according to whether they were run with PW_Coding_ or PW_LoF_ and whether the resulting model was significant (see Methods). NegCtrl are the same genes and phenotypes, but randomly re-assigned to pairs where there is no signal for association with the phenotype at the gene level. For quantitative traits, the stats given are the PW effect size divided by the whole gene effect size, displayed so that a value of 1.2 is shown as a 20% increase. For display purposes, instances where the whole-gene and PW had effect sizes in opposite directions (n=31 NegCtrl models) or where neither model produced an effect size >|0.5| (an additional n=4 NegCtrl models, n=12 non-significant models) are considered 0% improvement; the means shown in the graph therefore do not match those written in the text. Four models with >200% improvement are not shown (2 NegCtrl models, for *ANGPTL3* with mean corpuscular volume and *SLC22A12* with mean platelet volume; and 2 non-significant models, for *HBB* and *KLF1* with mean corpuscular volume). For binary traits, the stats given are the PW OR divided by the whole gene OR. For display purposes, instances where the ORs for the whole gene and PW models were in opposite directions (n=22 NegCtrl models, n=3 non-significant models) or where neither the whole-gene nor PW model produced an OR>2 (an additional n=12 NegCtrl models, n=8 non-significant models) are considered to have no improvement and thus a fold improvement of 1.

In contrast, the PW_LoF_ models show little to modest improvement over using the whole gene for either quantitative or binary traits and generally do not show a significant difference between PW and non-PW regions, with only a few exceptions (**Figure 3A & 3B** bottom**; S5B, S6B**). For most genes, when there is a LoF association, the statistical signal for LoF variants is spread across the entire gene, resulting in nearly the entire gene being included in PW_LoF_ models. This results in fewer significant PW_LoF_ models compared to PW_Coding_ models. For example, LoF variants anywhere in *LDLR* are generally considered pathogenic, and indeed PW_LoF_ implicates the entire gene (**Figure S6B**). However, when they are significant, PW_LoF_ models perform well: the average fold improvement for significant PW_LoF_ models over the whole gene model is 1.7x (range 1.2-2.9; binary traits) and change in effect size is 77% (range 10-160%; quantitative traits) (Figure 4). For example, the PW_LoF_ model for *TTN* with cardiomyopathy improves the OR by 2.9x in the test set by mostly restricting to cardiac-expressed exons, and the PW_LoF_ model for *APOB* with LDL levels produces an effect size improvement of 111% by removing the last 1082 bases of the gene (two-thirds of the final exon) as well as three single variants that did not show associations (**Table S1, S2**)^19^. We also test using LOFTEE to remove low confidence LoF variants from the whole gene models, which improves the effect size for some genes, but does not consistently result in a positive change (**Figure S5B, S6B**).

In contrast, our phenotypic negative control analysis does not show the same distinctions for separating the PW regions from the non-PW regions: all of the gene-disease associations have overlapping 95% CIs for the PW and non-PW models in the test set, indicating no significant PW models, as expected (**Figure S7**).

Finally, we test the significant PW models in an independent cohort of 28k individuals from the Healthy Nevada Project (HNP). In the 65 genes, there are a mean of 35 / 9 new Coding / LoF variants tested per gene in this set, as well as 63 / 9 Coding / LoF variants per gene that were already used in the UKB analysis (for *TTN*, which we summarize separately, these numbers are 1616 / 137 new and 2816 / 76 already used). We find that all 23 significant models with sufficient data available (see Methods) show the same direction of effect in this replication cohort, although due to the smaller sample sizes, the confidence intervals overlap between the PW and non-PW models in all but 6 of the models in the HNP (**Figure S8**). Testing the 26 non-significant PW models with sufficient data available shows 15 with the same direction of effect: additional studies with larger sample sizes will clarify which of these additional models are supported.

Overall, our results demonstrate that the PW methodology successfully segments gene regions that contribute to gene-based collapsing analysis signals, dramatically improving the effects seen compared to whole-gene models.

### Power Window distinguishes both regions and single variants that impact a trait

The crux of PW is a region-based analysis, which merges together the effects of variants that are too rare to drive a statistical signal on their own. However, with sample sizes in the hundreds of thousands, many rare variants (MAF<0.1%) still have enough carriers to achieve reasonable power for discovery on their own. To overcome the possibility of a single variant with many carriers overpowering the effects of nearby variants that are more rare, PW analyzes variants that are above the carrier cutoff, in this case 20 carriers (MAF 0.003%) and up to 600 carriers (MAF 0.1%), separately as their own unit instead of combining them into windows. We refer to these as single variant windows.

We examine to what extent the single variant windows contribute to our results, as opposed to the windows that combine multiple rare variants. We find that single variant windows constitute a mean of 56% / 36% (PW_Coding_ / PW_LoF_) of the carriers of qualifying variants in each gene. For the variants retained in the PW models, single variant windows constitute a mean of 34% / 28% of the PW carriers. In contrast, for the non-PW regions, single variant windows make up a mean of 59% / 56% of the carriers (**Figure S9**).

Classifying single variant windows as PW or non-PW makes a major contribution to most of the models tested. We build separate models that exclude the single variant windows, and we find that only 9 of the 22 significant quantitative models and 2 of the 7 significant binary models retain significance using the same criteria as above. As a negative genetic control, we randomly assign locations for the rare variants within each gene and build new models. We find that only 1 of the models remains significant with this negative control: *TUBB1* with platelet distribution width. In this case, the original model without single variant windows retains 75% of the carriers in the gene, meaning that changing the locations of the variants would still implicate most of the gene, and some of the non-associated regions include variants with >10 carriers, which have high enough frequencies to dampen the signals of their newly assigned windows and cause the effect size from the non-PW carriers to have non-overlapping 95%CIs with those of the PW carriers.

Assessing the effect of each variant individually is the ideal scenario for association studies. As sample sizes grow, more and more rare variants will have sufficient carriers to be analyzed individually instead of grouped together. By focusing on effect size instead of p-value, our method only incorporates single variants into the model when they are having as big of an impact on an individual’s phenotype as do groups of rare variants for that gene.

### Power Window successfully isolates gene regions with different directions of effect

Each of the associations that are refined with PW has an underlying story that is rooted in the biology of the gene and the phenotype. One unique refinement that PW can make over a whole-gene model is identifying regions of the gene with opposite directions of effect, especially for quantitative traits. PW models identify opposite directions of effect within the same gene for 8 PW_coding_ models and 1 PW_LoF_ model (**Figure S6**). However, only two were part of significant PW models in the test set (**Figure 3A**).

One significant PW_coding_ model with opposite directions of effect is *GCK*, where we identify that non-benign coding variants toward the 5’ end of the gene are associated with low glucose levels, while coding variants in most of the rest of the gene are associated with high glucose levels (**Figure 1)**. Intriguingly, two variants in these models that are single variant windows (rs373418736 and rs143484733), meaning that they are rare (MAF<0.1%) yet common enough to be analyzed separately (n>20 carriers), show opposite directions of effect as compared to the rest of the region. The PW methodology of breaking out single variant windows allows these opposing signals within the same regional location to be separated out and analyzed appropriately.

The other significant PW_coding_ model with opposite directions of effect is *LDLR* with LDL levels. Here, we identify that non-benign coding variants toward the 3’ end of the gene, corresponding to the transmembrane and cytoplasmic portions, are associated with lower LDL levels, while non-benign coding variants in most of the rest of the gene, which would be extracellular and thus where LDL binds to this gene product, are associated with higher LDL levels. This is consistent with the mechanism of disease, where the LDLR receptor no longer interacts/binds LDL well, leaving cholesterol in the blood stream which results in high measured blood biomarker levels). The coding variants associated with the higher LDL levels in our dataset are also associated with coronary artery disease to the same level as known coding P/LP variants from ClinVar (**Figure S10**).

## Discussion

Here, we present a new method called Power Window (PW) that successfully identifies the variants and regions within genes that are responsible for the statistical associations found in gene-based collapsing analyses of rare variants. We tested this new method on 65 established gene-phenotype associations of rare variants collapsed at the gene level. Our method distinguishes between the variants and gene regions that do and do not impact these traits, identifying statistically significant differences between distinct regions of the gene 46% of the time for quantitative traits with a coding model, and ∼15% of the time for binary traits and LoF models. The method often dramatically improved the association signals, for example improving the normalized effect size by a mean of 40% for significant coding models for quantitative traits when compared to an analysis that collapses across the whole gene. PW also identifies when the entire gene is implicated instead of specific regions, which was true for 52% of PW_LoF_ models and 3% of PW_coding_. Finally, in some cases, PW even identifies variants and regions with opposite directions of effect in the same gene.

PW uses the concept of a sliding window to distinguish gene regions that are associated with a phenotype from those that are not. However, unlike sliding window approaches that move according to a fixed number of bases or variants, PW slides according to the most important metric for rare variant analyses: the statistical power. This focus on power, in this case dictated by the number of individuals carrying rare qualifying variants within a given region, is the critical feature for its success and is a key feature of its flexibility. Without this crucial innovation, it is difficult to tell whether regions without statistical associations are truly not associated or simply lack power. Performing an analysis where all the windows of the gene have the same statistical power allows you to rank the parts of the gene according to their associations in a manner that is unbiased.

Performing discovery analyses with PW is only now feasible with the availability of very large sample sizes from large population studies and biobanks and will continue to improve as sample sizes increase. Being able to break a gene down into windows and still have a reasonable number of carriers of rare genetic variants–which are themselves rare events–in each window requires sample sizes that until now had been unreasonable, and which remain unreasonable for many genes. For example, *MIP*, which has a significant association with cataracts, had 52 carriers of qualifying variants in the set of 300k UKB training exomes. A window size of 20 carriers produces only 51 windows for this gene, making it much less practical to study than a gene like *TTN*, where 1,349 windows were made for the LoF model.

PW is able to extend known LoF associations to coding variants by identifying the regions of the gene in which coding variants have an effect similar to that of LoF variants. This is important because the complexity of interpreting novel coding variants has remained a difficult problem in human genetics. For example, ACMG guidelines allow novel LoF variants to be considered pathogenic in a gene where other LoF variants are already established as pathogenic, whereas each individual non-LoF variant requires, for example, evidence of association in other patients and functional studies to support their pathogenicity^28^. PW uses statistical evidence to distinguish coding variants, even novel ones, that have an impact similar to those of LoF variants. As an example, LoF variants in *LDLR* are considered pathogenic^24^. Indeed, our PW_LoF_ model confirms that LoF variants anywhere in this gene are associated with higher LDL levels (effect size 1.2; **Figure S6B**). However, PW_coding_ identifies that 45% of the carriers of rare non-benign coding variants in this gene are also associated with higher LDL levels (effect size 0.6, **Figure 3**). In this way, PW can uncover latent non-LoF signals from regions of a gene that might otherwise go unobserved. This extension of known biology to additional types of variants in the gene increases the number of people who would benefit from genetic screening and also improves our understanding of gene function.

PW was able to identify when variants in different gene regions could produce significantly different directions of effect on a phenotype. Among these was the association between *GCK* and glucose levels. It is known that depending on the variant effect on overall glucose metabolism, mutations can lead to either hyper- or hypo-glycemia^25^. We find both of these effects, concurrently, with the PW analysis, highlighting the regions of the gene structurally and functionally associated with each resulting phenotype. While hypoglycemic variants have been reported throughout various regions of *GCK*, we find most variants are concentrated toward the 5’ end, some of which are very well described while others may help to further our mechanistic understanding of *GCK* and thus improve the scope of variant interpretation possible at this locus^25^. This finding is also supported by a recent deep scan mutagenesis study which found that 5’-end mutations were more likely to lead to lower glucose levels.^26^.

While the theoretical basis for the PW method is a substantial advancement for identifying the specific regions of genes in which rare variants are associated with traits, there are still many refinements to the methodology that will be beneficial for future studies. One aspect is that the method requires a balance between zooming in on specific regions of the gene (requiring a small carrier sample size cutoff) and obtaining statistical significance for the regions (requiring a larger carrier sample size cutoff). Future studies with even larger sample sizes may reclassify some portions of the genes tested here, identifying some regions we erroneously excluded or identifying new regions to include. Other improvements will involve focusing on certain classes of variants in the gene, such as those computationally predicted to be gain or loss of function, those with functional data from screenings, and those in transcripts expressed in tissues of interest^19,26,27^.

The PW method is centered on statistics and can accommodate all classes of genetic variation to fine-tune rare variant signals and gene-based results. We believe the PW technique can have an immediate impact on human genomics research, drug development, variant interpretation and precision medicine. As displayed in this work, PW can both improve the specificity of variant interpretation for rare variants in established gene-level disease associations through regional refinement of the association. It can also expand variant classification capabilities by combining PW analyses derived from the same phenotype with carriers defined by both LoF and coding variant models. In both cases, with more precise or complete genomic definitions to identify relevant carriers, phenotypic penetrance estimates amongst carriers are likely to improve– a key metric for the practice of precision medicine. We also anticipate that PW will be a powerful tool for discovery, as it will be able to identify sub-gene-disease associations that are drowned out when rare variants are collapsed at the gene level, characterize variant patterns in a locus to better understand the mechanism of disease, and improve knowledge of gene function to predict new drug targets.

## Data Availability

Statistics relating to the Power Window analysis for all included genes, calculated using the UKB 300K discovery cohort, are available in Table S3. UKB data are available for download (https://www.ukbiobank.ac.uk/) to qualified researchers. The HNP data are available to qualified researchers upon reasonable request and with permission of the Institute for Health Innovation (IHI) and Helix. Researchers who would like to obtain the raw genotype data related to this study will be presented with a data user agreement which requires that no participants will be re-identified and no data will be shared between individuals or uploaded onto public domains. The IHI encourages and collaborates with scientific researchers on an individual basis. Examples of restrictions that will be considered in requests to data access include but are not limited to (1) whether the request comes from an academic institution in good standing and will collaborate with our team to protect the privacy of the participants and the security of the data requested, (2) type and amount of data requested, (3) feasibility of the research suggested, (4) amount of resource allocation for the IHI and Renown Hospital required to support the collaboration. Any correspondence and data availability requests related to HNP should be addressed to JG at or Craig Kugler.

## Acknowledgements

This research has been conducted using the UK Biobank Resource under Application Number 40436. Funding was provided to DRI by the Nevada Governor’s Office of Economic Development. Funding was provided to the Renown Institute for Health Innovation by Renown Health and the Renown Health Foundation. We acknowledge the entire Helix Bioinformatics team for their contributions to the production exome sequencing pipeline. We thank all of the genomic representatives of the Healthy Nevada Project (HNP). We thank Renown Health and DRI marketing for helping to launch the HNP.

## Data code and availability

Statistics relating to the Power Window analysis for all included genes, calculated using the UKB 300K discovery cohort, are available in **Table S3**. UKB data are available for download (https://www.ukbiobank.ac.uk/) to qualified researchers. The HNP data are available to qualified researchers upon reasonable request and with permission of the Institute for Health Innovation (IHI) and Helix. Researchers who would like to obtain the raw genotype data related to this study will be presented with a data user agreement which requires that no participants will be re-identified and no data will be shared between individuals or uploaded onto public domains.

The IHI encourages and collaborates with scientific researchers on an individual basis. Examples of restrictions that will be considered in requests to data access include but are not limited to (1) whether the request comes from an academic institution in good standing and will collaborate with our team to protect the privacy of the participants and the security of the data requested, (2) type and amount of data requested, (3) feasibility of the research suggested, (4) amount of resource allocation for the IHI and Renown Hospital required to support the collaboration. Any correspondence and data availability requests related to HNP should be addressed to JG at or Craig Kugler.

