## Supplementary Figures for "A power-based sliding window approach to evaluate the clinical impact of rare genetic variants"

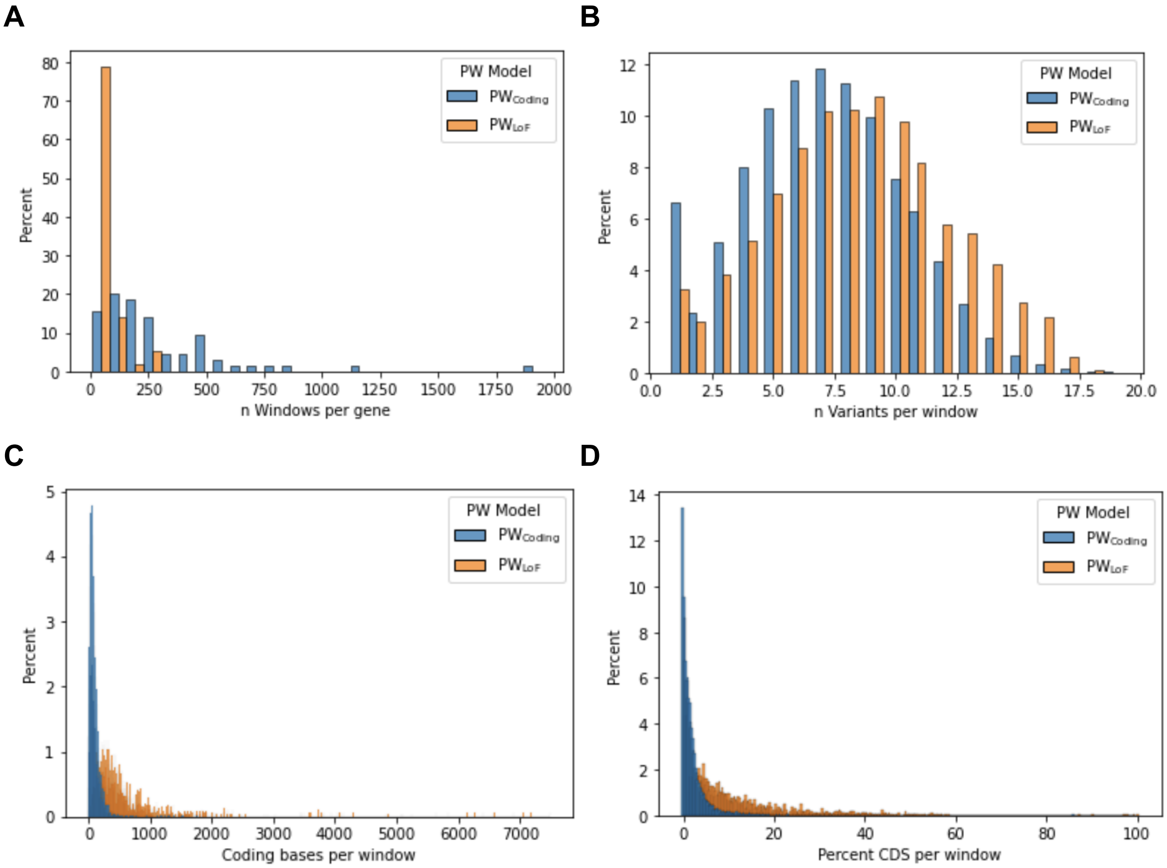


**Figure S1. Stats for power windows.** Because of the size of *TTN*, the stats for this gene are not displayed and instead are written separately in the legend. A) Histogram of number of windows analyzed per gene. The mean number of windows analyzed per gene was 304 / 66 for $PW_{Coding}$ / $PW_{LoF}$ (range 35-1939 and 7-300). *TTN* = 13,672 / 1349 for $PW_{Coding}$ / $PW_{LoF}$. B) Histogram of number of variants included in each window. The mean number of variants analyzed per window was 7 / 9 for $PW_{Coding}$ / $PW_{LoF}$ (range 1-19 and 1-18). *TTN* = 7 (1-18) / 11 (1-18) for $PW_{Coding}$ / $PW_{LoF}$. C) Histogram of number of coding bases included in each window; when a window only included one variant or only included variants at the same site, then the length is 1. The mean number of coding bases included per window was 76 / 429 for $PW_{Coding}$ / $PW_{LoF}$ (range 1-1774 and 1-7047). *TTN* = 58 (1-736) / 1111 (1-3496) for $PW_{Coding}$ / $PW_{LoF}$. D) Histogram of the percent of each CDS for the gene that was included in each window. The mean percent of the CDS of the gene that was analyzed in each window was 2% / 11% for $PW_{Coding}$ / $PW_{LoF}$ (range <1-86% and <1->99%). *TTN* = 0.05% (0.001%-0.7%) / 1.0% (0.001%-3.2%) for $PW_{Coding}$ / $PW_{LoF}$.

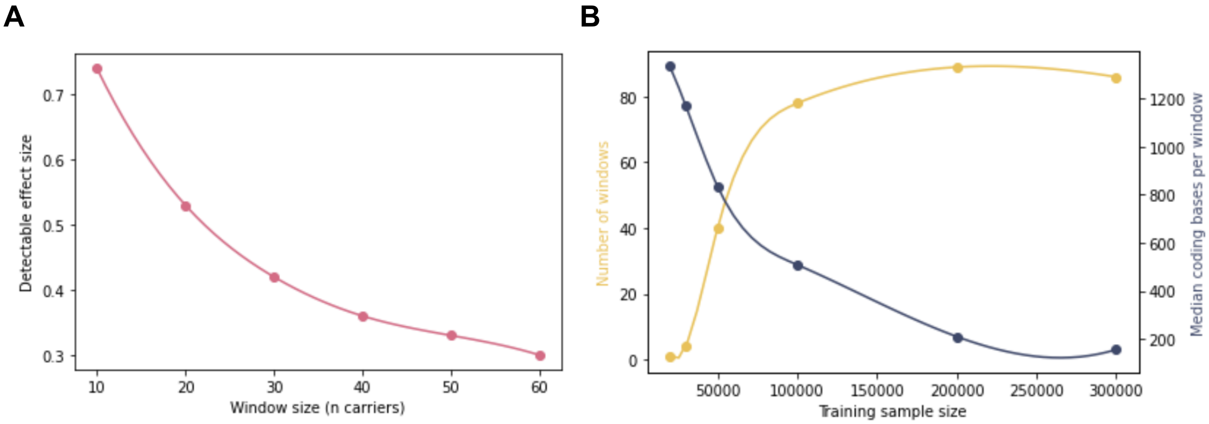


**Figure S2. Properties of Power Window size**. A) For a quantitative trait in a training set of 300k samples, the window size (number of carriers in the window) is compared to the effect size for which there is power to identify (with 99% confidence) that the beta is not 0. B) For the example gene *GCK* with a coding model (which in our main model had 86 windows, with a mean window length of 146 coding bases), the training sample size is compared to the physical length of each window in terms of coding position, for a set window size of 20 carriers per window. A larger sample size results in being able to home in on more specific regions for analysis.

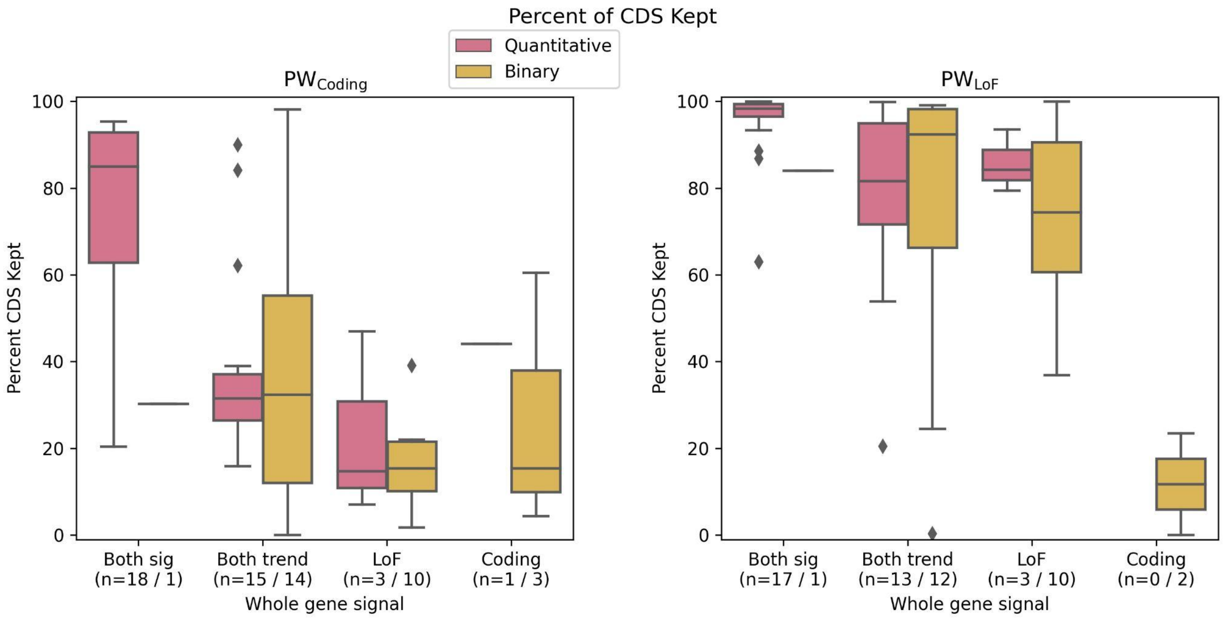


**Figure S3. Percent CDS kept in Power Window models.** For each gene (n=65), the percent of the CDS for the gene that was retained by the PW model was evaluated for (A) $PW_{Coding}$ and (B) $PW_{LoF}$. Within each model, the phenotypes were grouped into quantitative (pink) or binary (yellow) traits, and the genes were grouped based on the original genome-wide association as follows: **both sig**: original associations were genome-wide significant for both a coding and LoF models (p<5e-8); **both trend**: genome-wide significance was shown for either coding or LoF, the other model showed at least a nominal association (p<0.05); **LoF**: LoF only and not coding; **coding**: coding only and not LoF. For all genes, at least one window was included in the final Power Window (PW) model, with the exception of *JAK2* with myeloproliferative disease for the LoF model, which had whole-gene association signal only for the coding model. Axis label indicates the number of genes tested within each group.

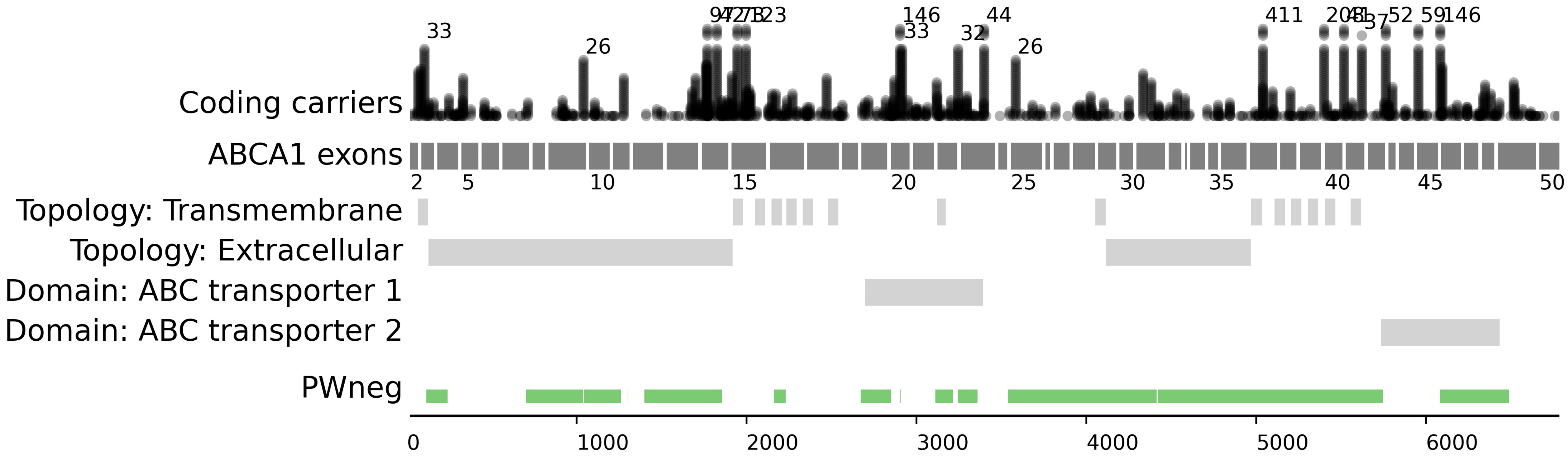
Coding carriers
ABCA1 exons
Topology: Transmembrane
Topology: Extracellular
Domain: ABC transporter 1
Domain: ABC transporter 2
PWneg
33
26
146
33
32
44
26
411
37
52
146
2
5
10
15
20
25
30
35
40
45
50
0
1000
2000
3000
4000
5000
6000

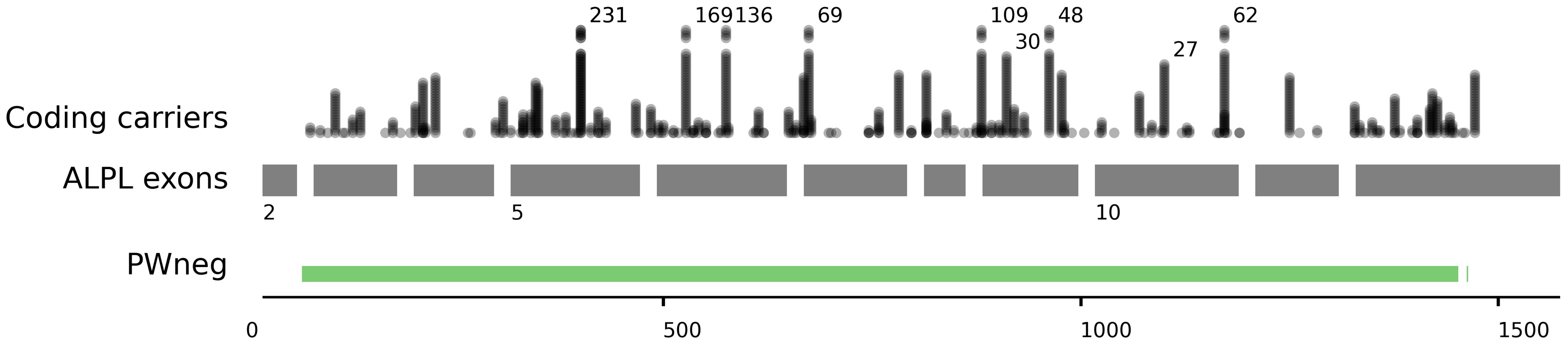

Coding carriers
ALPL exons
PWneg
231
169136
69
109
30
48
27
62
2
5
10
0
500
1000
1500

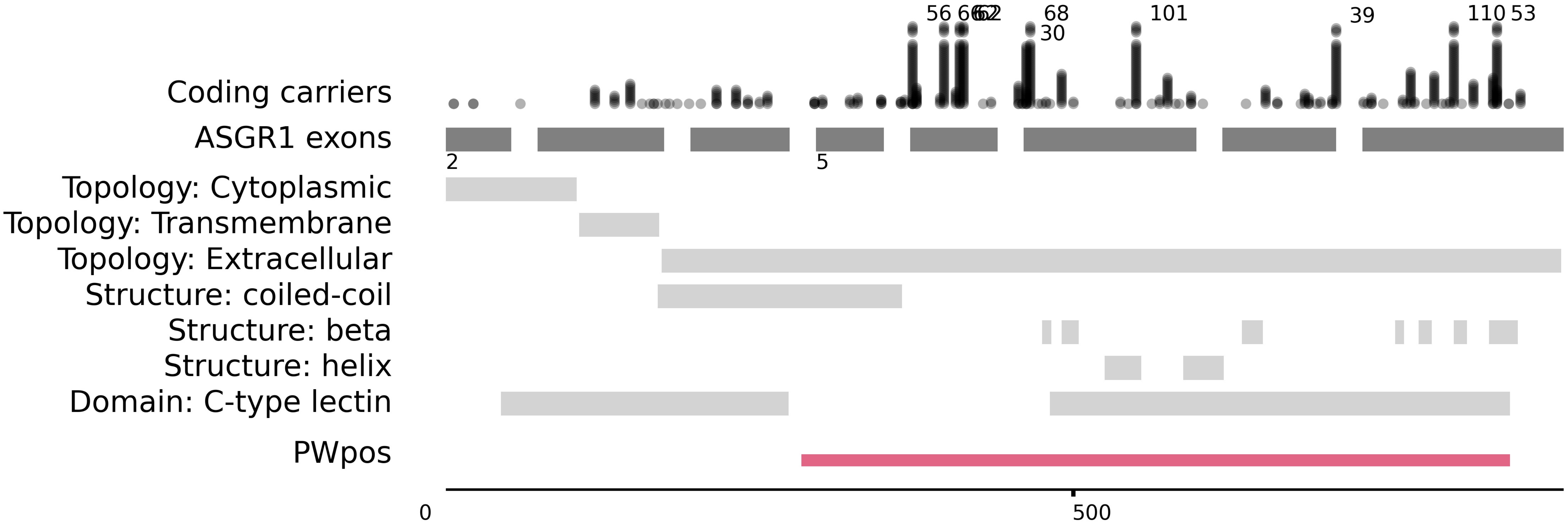
Coding carriers
ASGR1 exons
Topology: Cytoplasmic
Topology: Transmembrane
Topology: Extracellular
Structure: coiled-coil
Structure: beta
Structure: helix
Domain: C-type lectin
PWpos
56
68
30
101
39
110
53
2
5
0
500

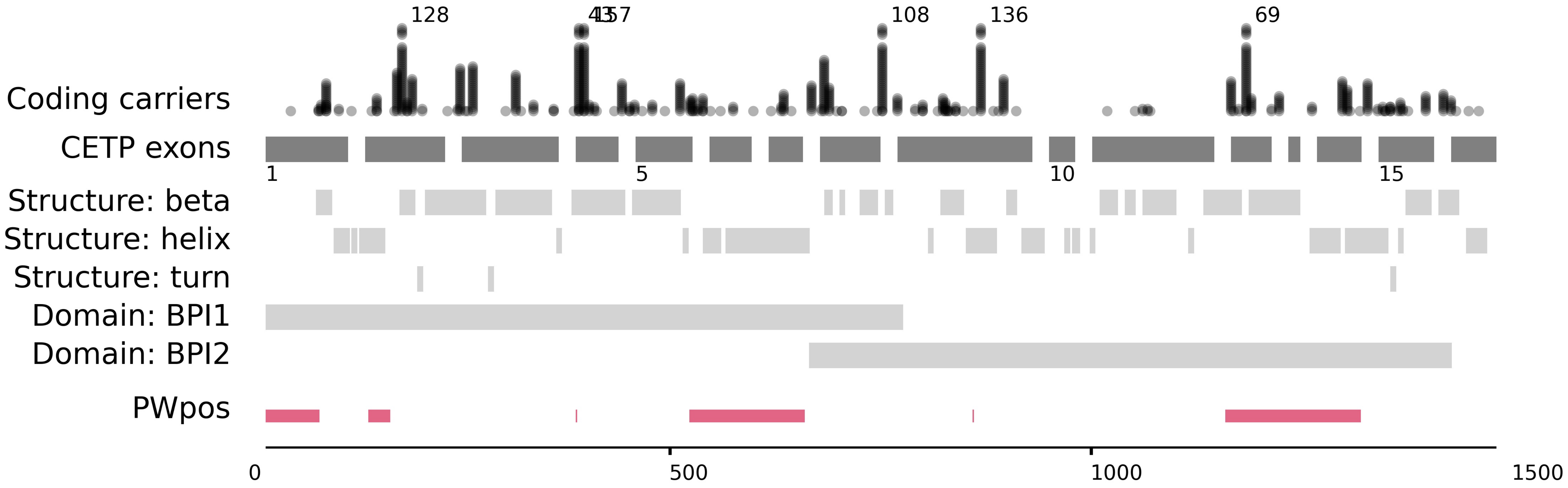

Coding carriers
128
4157
108
136
69
CETP exons
1
5
10
15
Structure: beta
Structure: helix
Structure: turn
Domain: BPI1
Domain: BPI2
PWpos
0
500
1000
1500

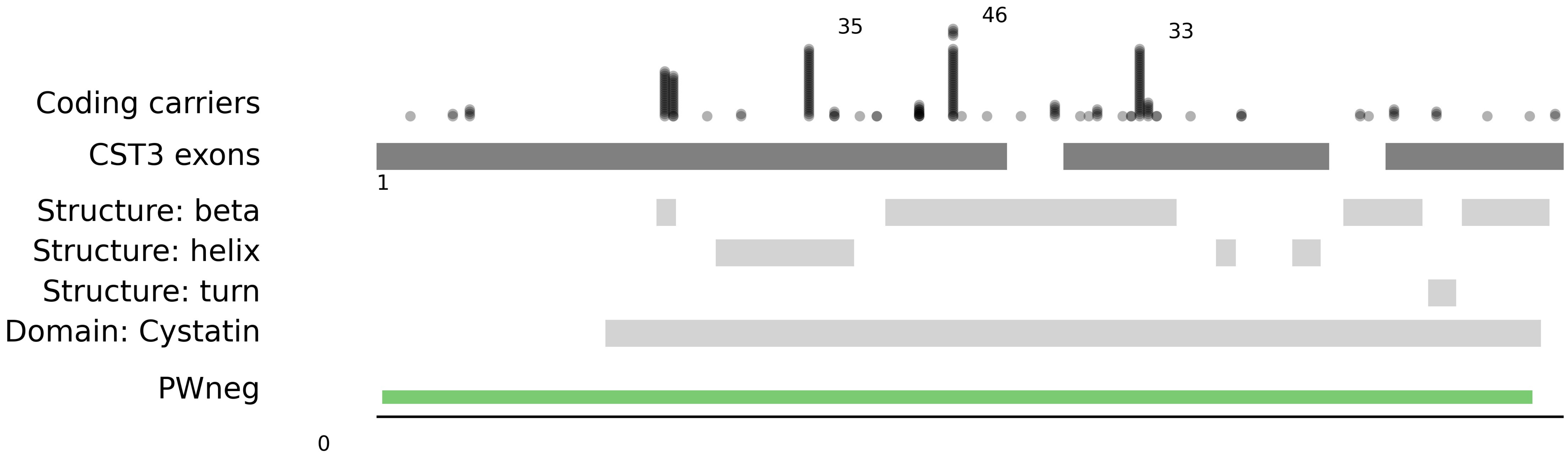

35
46
33
Coding carriers
CST3 exons
1
Structure: beta
Structure: helix
Structure: turn
Domain: Cystatin
PWneg
0

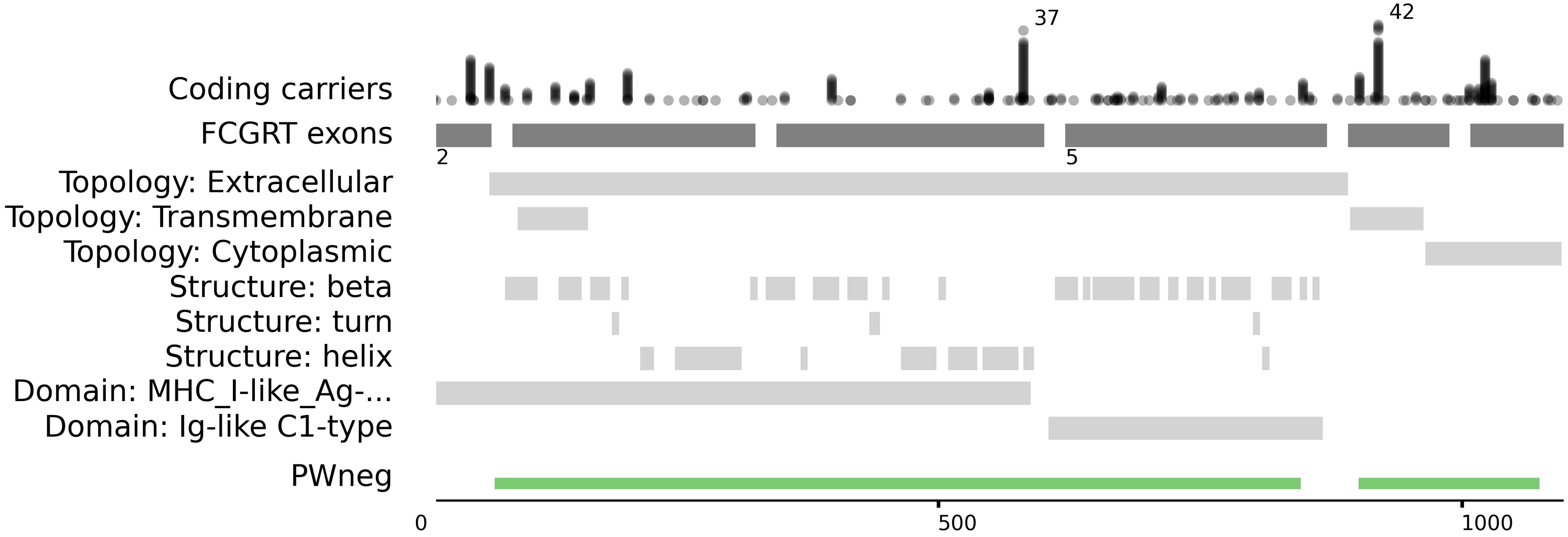

37
42
Coding carriers
FCGRT exons
2
5
Topology: Extracellular
Topology: Transmembrane
Topology: Cytoplasmic
Structure: beta
Structure: turn
Structure: helix
Domain: MHC_I-like_Ag-...
Domain: Ig-like C1-type
PWneg
0
500
1000

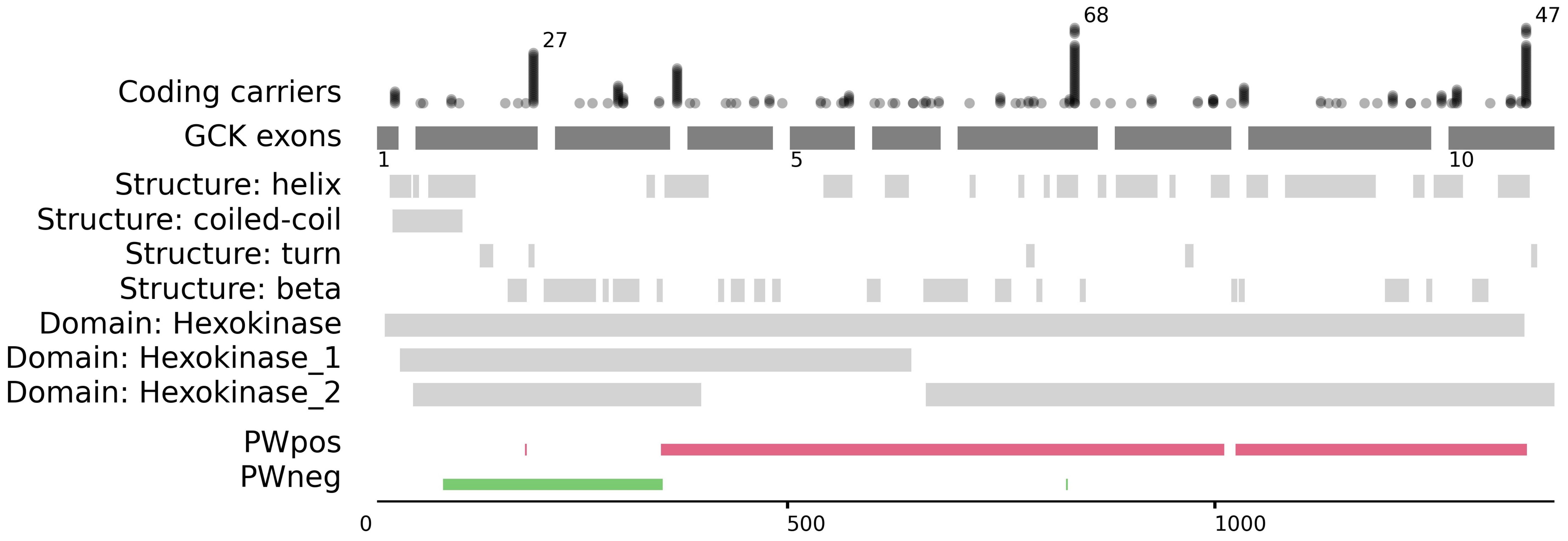

Coding carriers
GCK exons
Structure: helix
Structure: coiled-coil
Structure: turn
Structure: beta
Domain: Hexokinase
Domain: Hexokinase_1
Domain: Hexokinase_2
PWpos
PWneg
27
68
47
1
5
10
0
500
1000

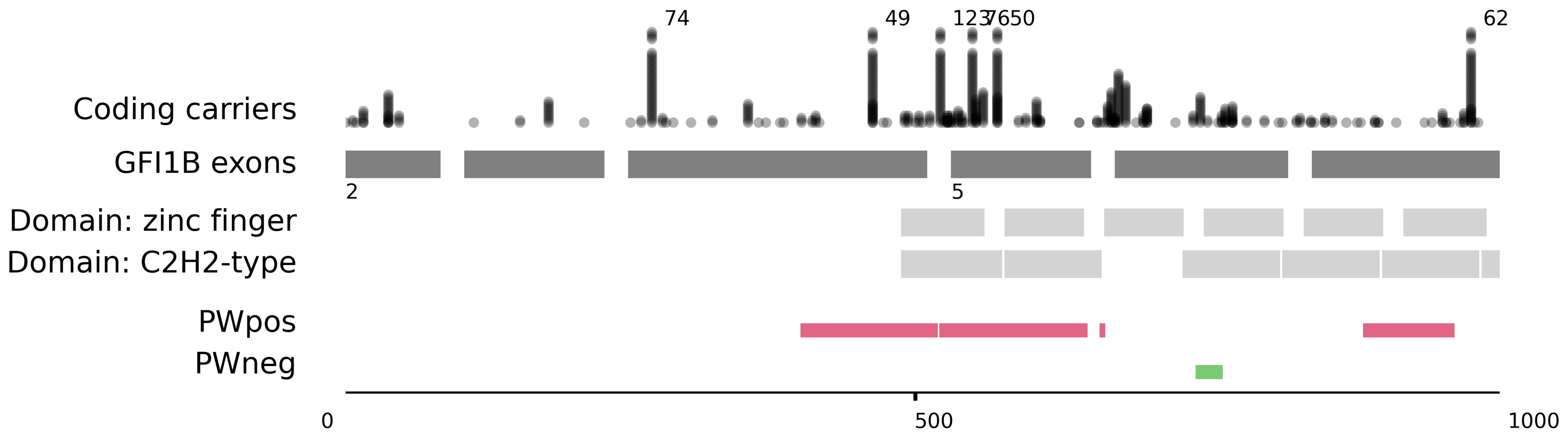
74
49
1237650
62
Coding carriers
GFI1B exons
2
5
Domain: zinc finger
Domain: C2H2-type
PWpos
PWneg
0
500
1000

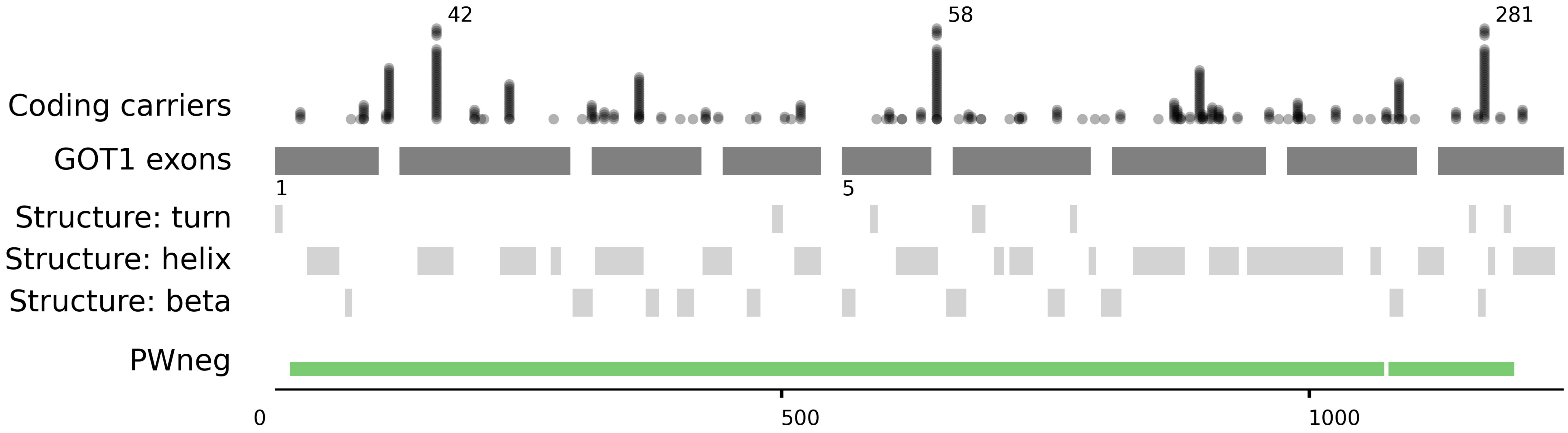

42
58
281
Coding carriers
GOT1 exons
1
5
Structure: turn
Structure: helix
Structure: beta
PWneg
0
500
1000

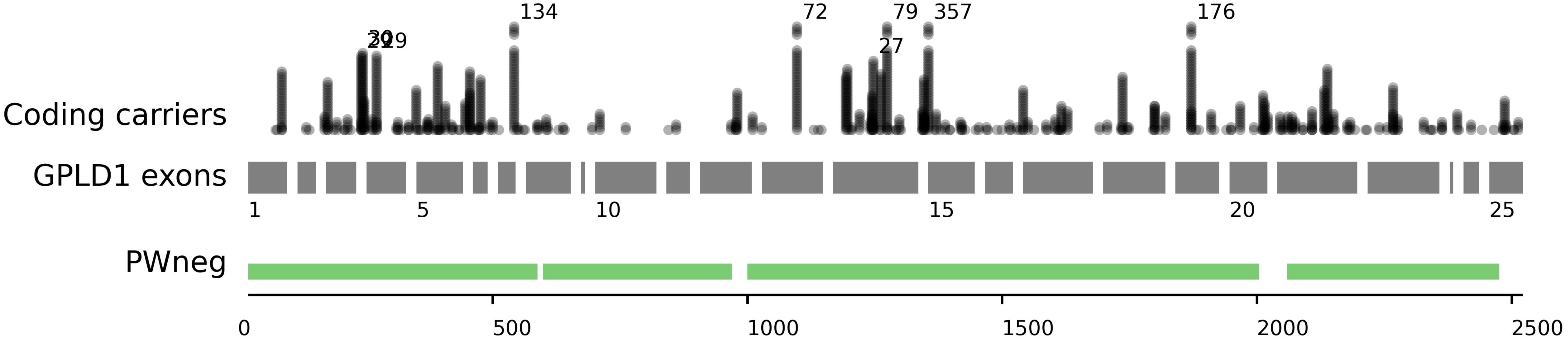

Coding carriers
GPLD1 exons
PWneg
134
72
79
357
176
27
1
5
10
15
20
25
0
500
1000
1500
2000
2500

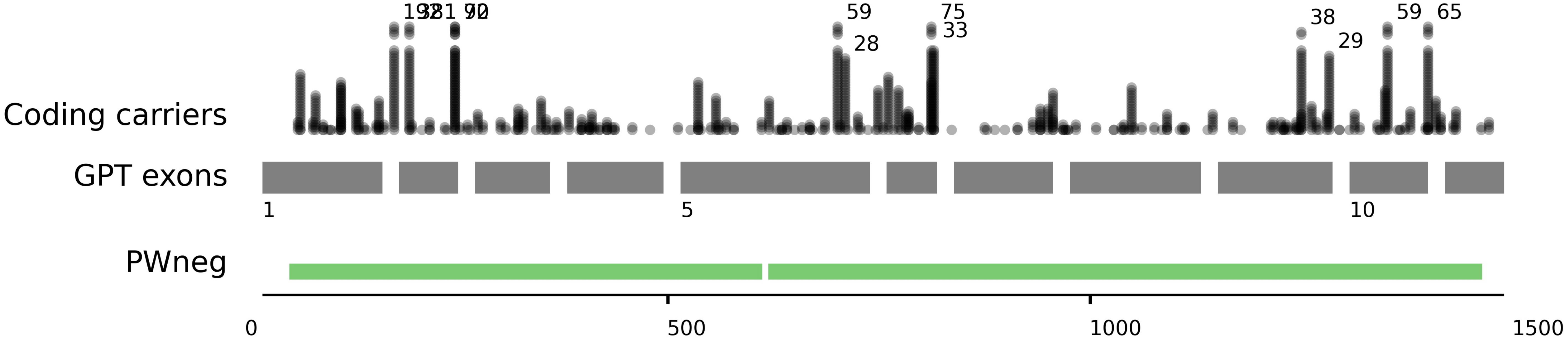

59
28
75
33
38
29
59
65
Coding carriers
GPT exons
1
5
10
PWneg
0
500
1000
1500

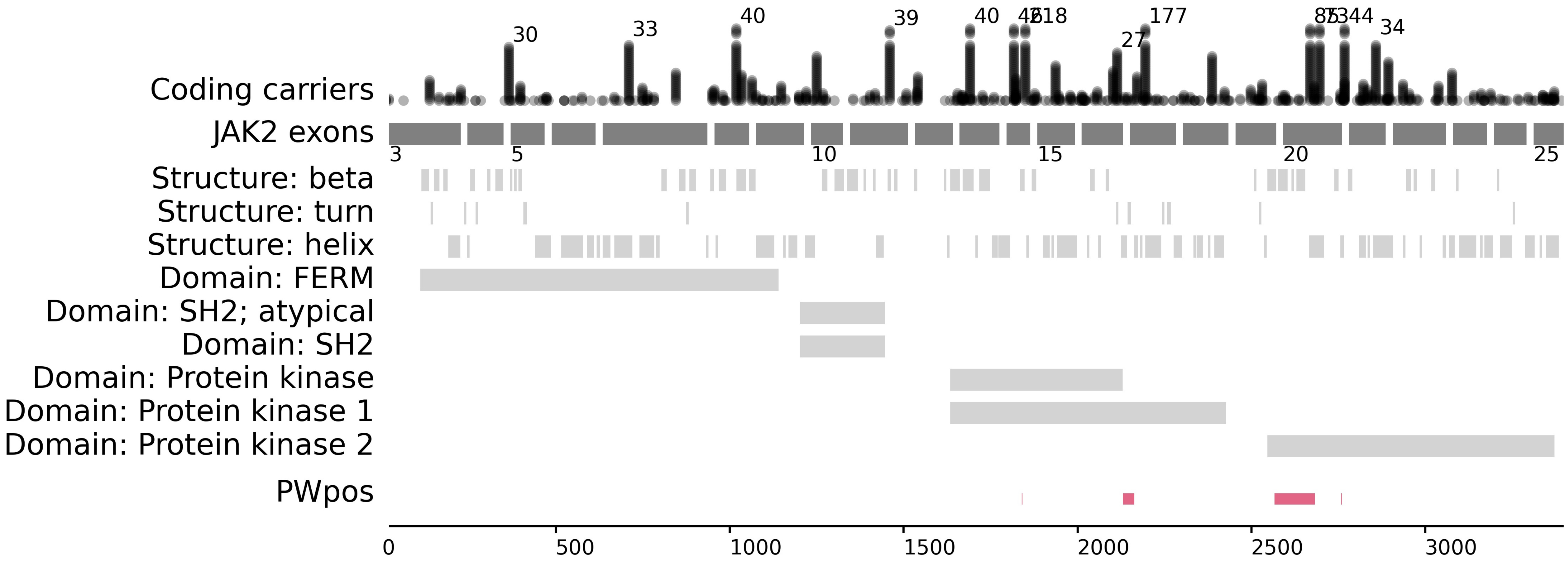
Coding carriers
JAK2 exons
Structure: beta
Structure: turn
Structure: helix
Domain: FERM
Domain: SH2; atypical
Domain: SH2
Domain: Protein kinase
Domain: Protein kinase 1
Domain: Protein kinase 2
PWpos
0
500
1000
1500
2000
2500
3000

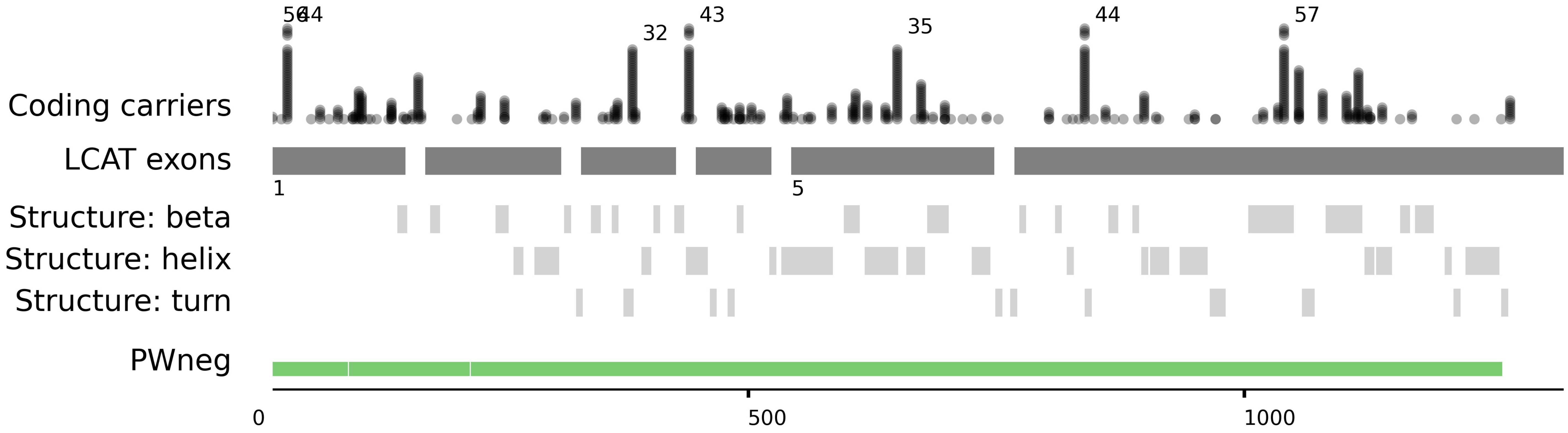

56
44
32
43
35
44
57
Coding carriers
LCAT exons
1
5
Structure: beta
Structure: helix
Structure: turn
PWneg
0
500
1000

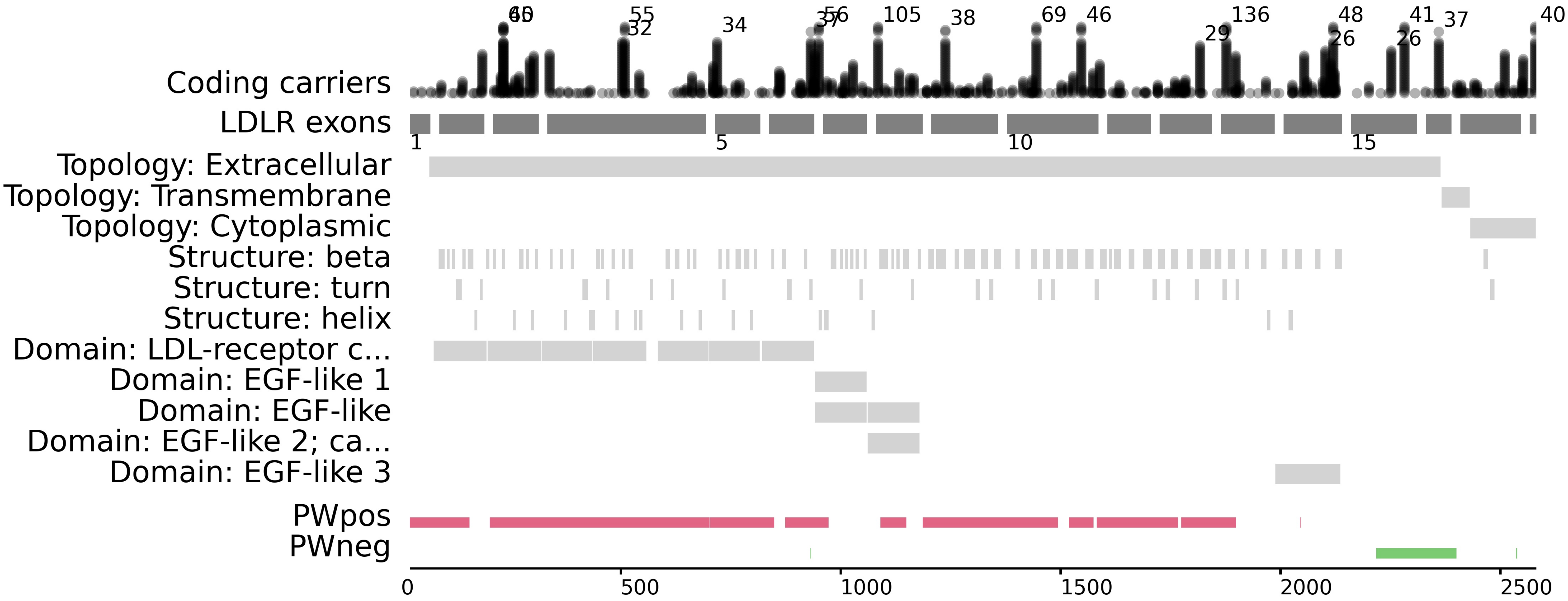
Coding carriers
LDLR exons
Topology: Extracellular
Topology: Transmembrane
Topology: Cytoplasmic
Structure: beta
Structure: turn
Structure: helix
Domain: LDL-receptor c...
Domain: EGF-like 1
Domain: EGF-like
Domain: EGF-like 2; ca...
Domain: EGF-like 3
PWpos
PWneg
1
5
10
15
0
500
1000
1500
2000
2500
55
32
34
105
38
69
46
29
136
48
26
41
26
37
40

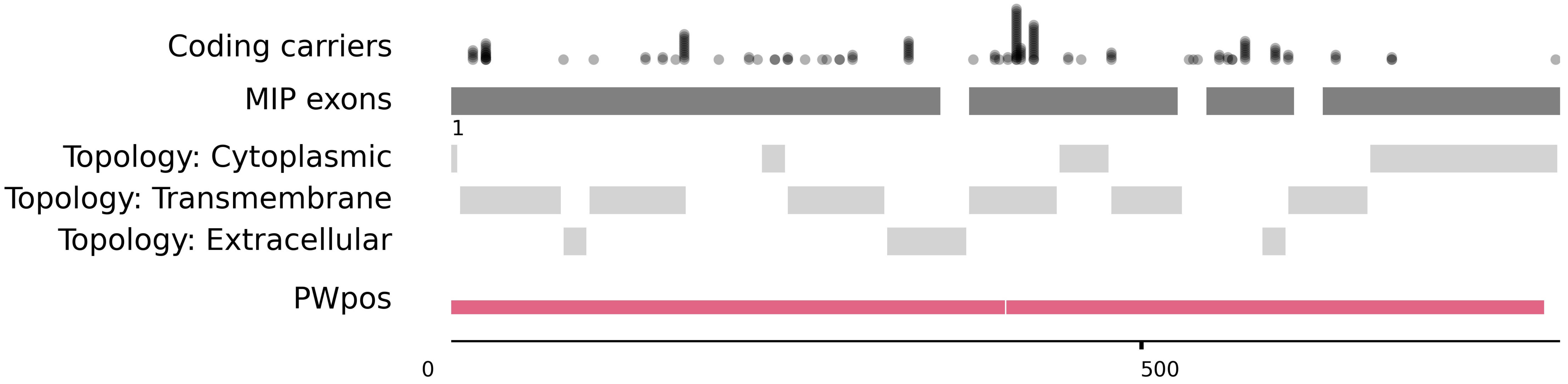

Coding carriers
MIP exons
1
Topology: Cytoplasmic
Topology: Transmembrane
Topology: Extracellular
PWpos
0
500

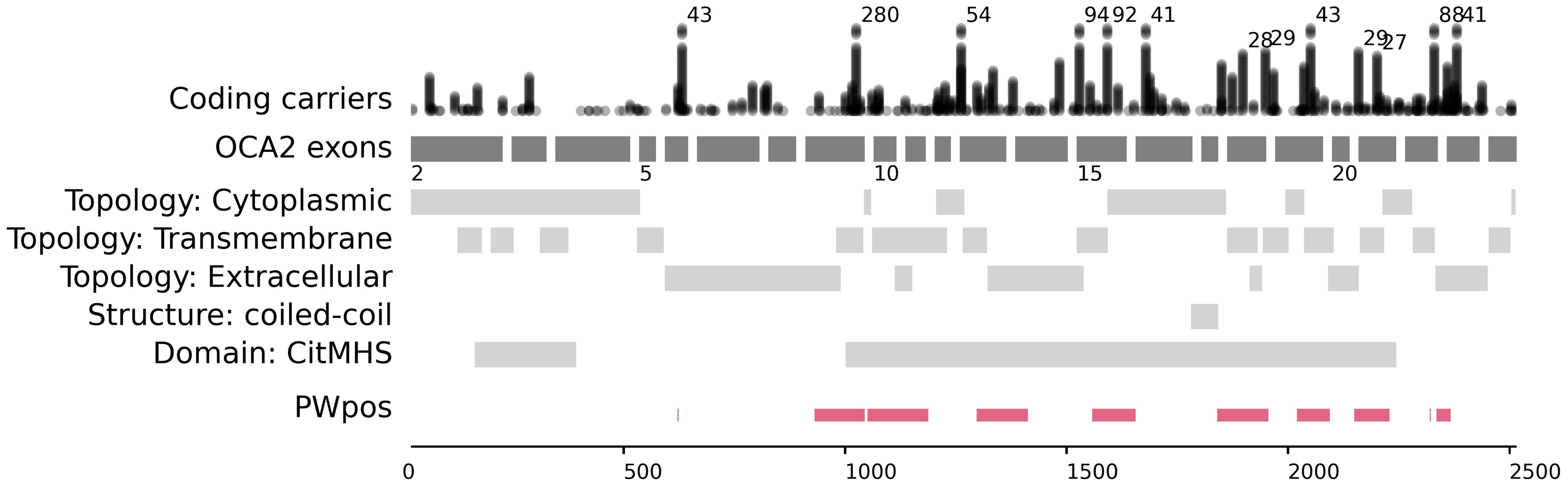

Coding carriers
OCA2 exons
Topology: Cytoplasmic
Topology: Transmembrane
Topology: Extracellular
Structure: coiled-coil
Domain: CitMHS
PWpos
43
280
54
94
92
41
28
29
43
29
27
88
41
2
5
10
15
20
0
500
1000
1500
2000
2500

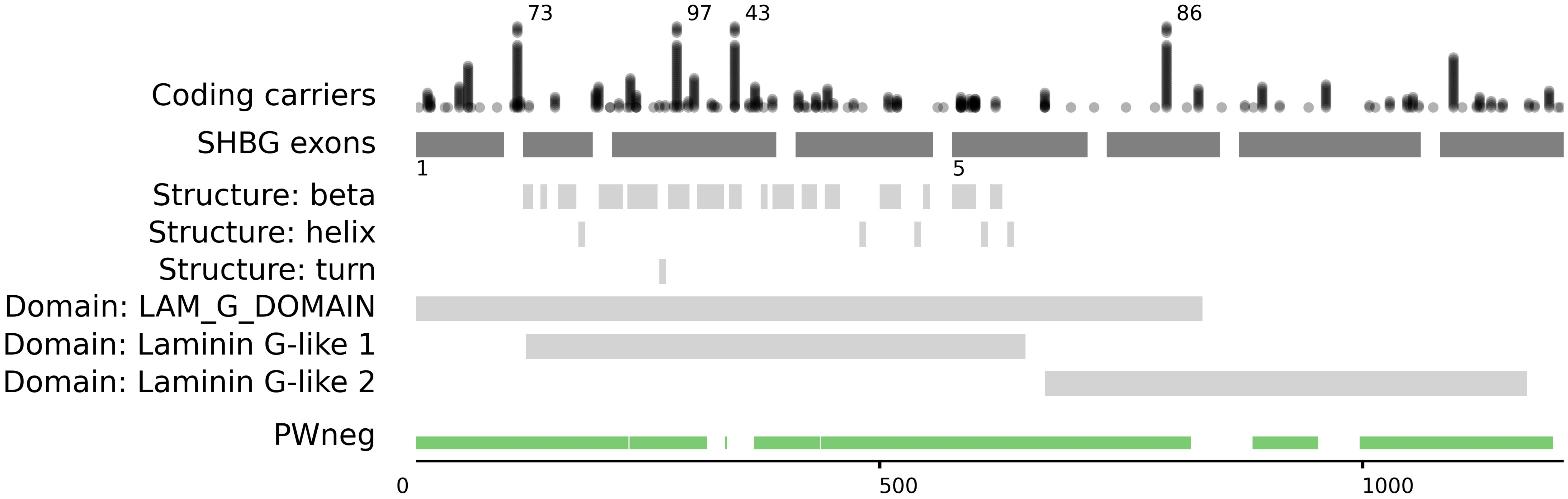

73
97
43
86
Coding carriers
SHBG exons
1
5
Structure: beta
Structure: helix
Structure: turn
Domain: LAM_G_DOMAIN
Domain: Laminin G-like 1
Domain: Laminin G-like 2
PWneg
0
500
1000

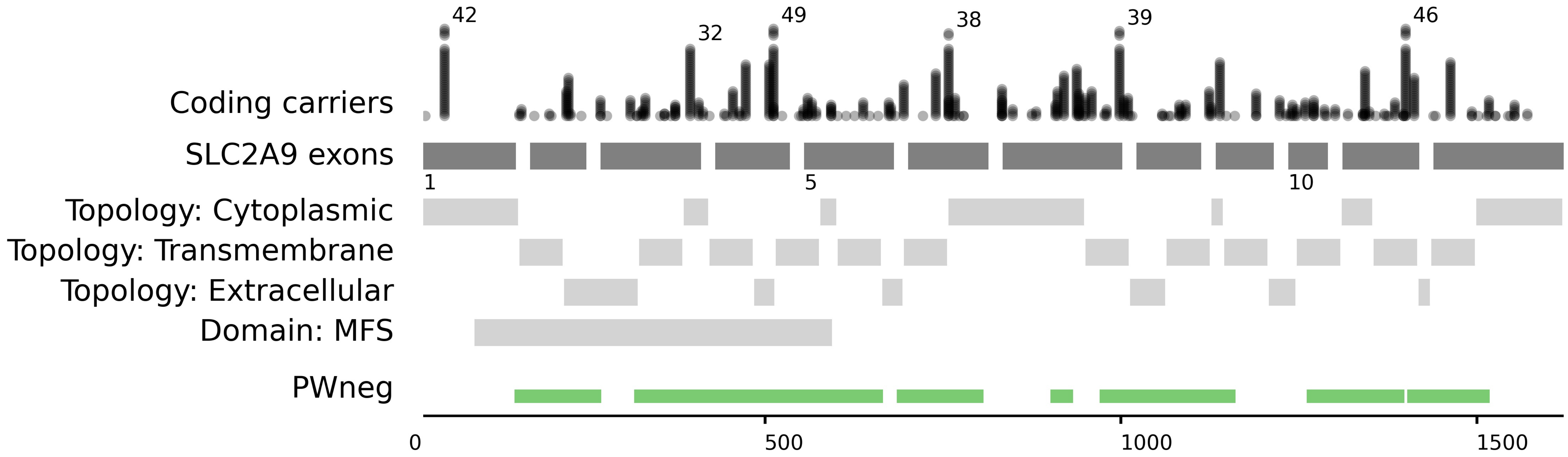

42
32
49
38
39
46
Coding carriers
SLC2A9 exons
1
5
10
Topology: Cytoplasmic
Topology: Transmembrane
Topology: Extracellular
Domain: MFS
PWneg
0
500
1000
1500

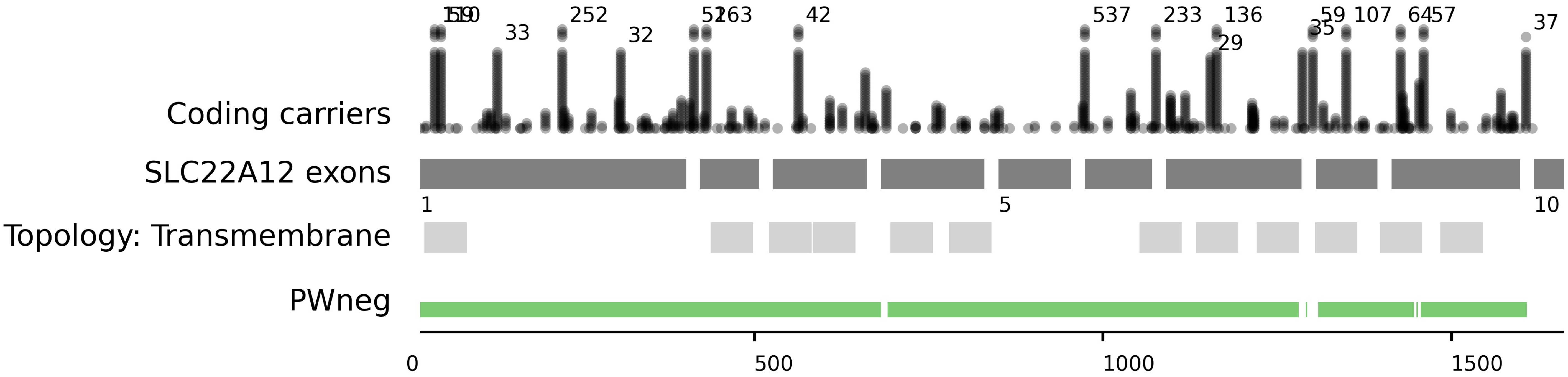

Coding carriers
SLC22A12 exons
Topology: Transmembrane
PWneg
1
5
10
0
500
1000
1500
33
252
32
42
537
233
136
29
35
107
37

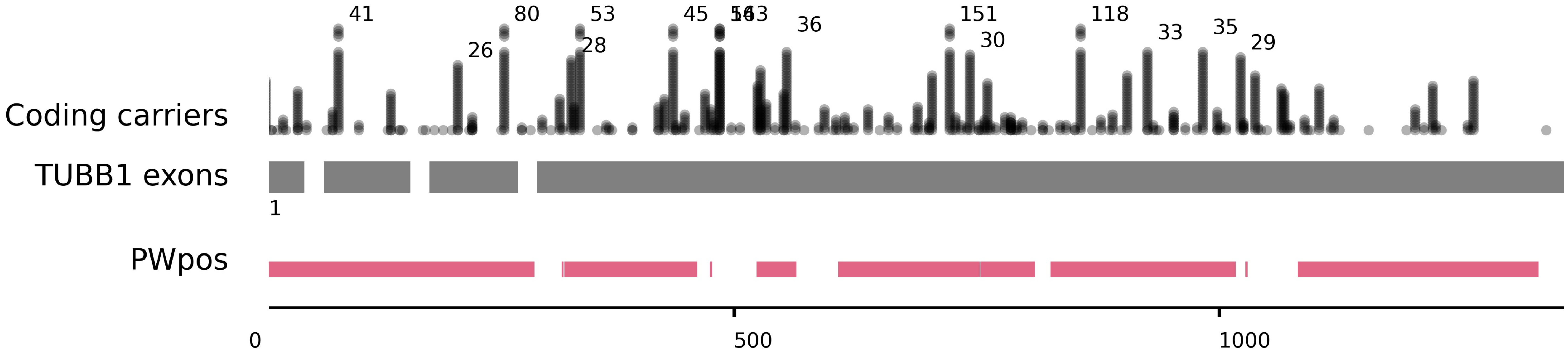

41
80
53
45
5163
36
151
118
33
35
29
26
28
30
Coding carriers
TUBB1 exons
1
PWpos
0
500
1000

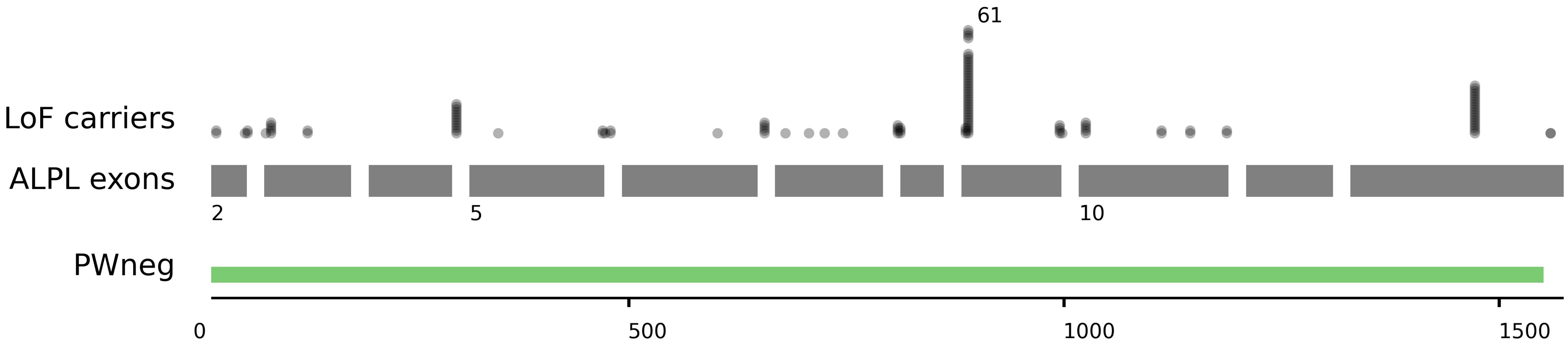

61
LoF carriers
ALPL exons
2
5
10
PWneg
0
500
1000
1500

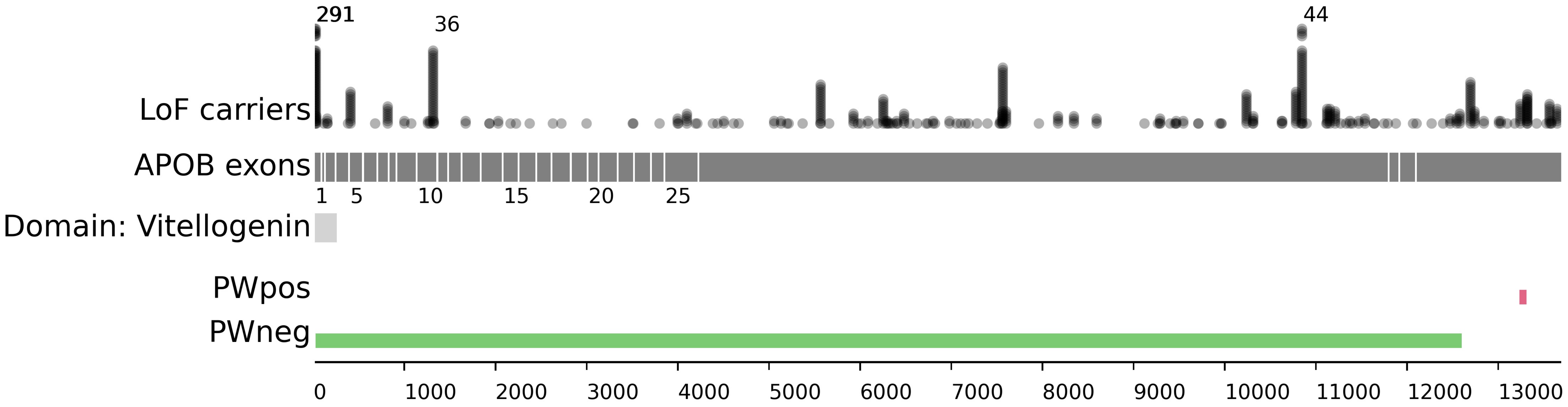

291
36
44
LoF carriers
APOB exons
1
5
10
15
20
25
Domain: Vitellogenin
PWpos
PWneg
0
1000
2000
3000
4000
5000
6000
7000
8000
9000
10000
11000
12000
13000

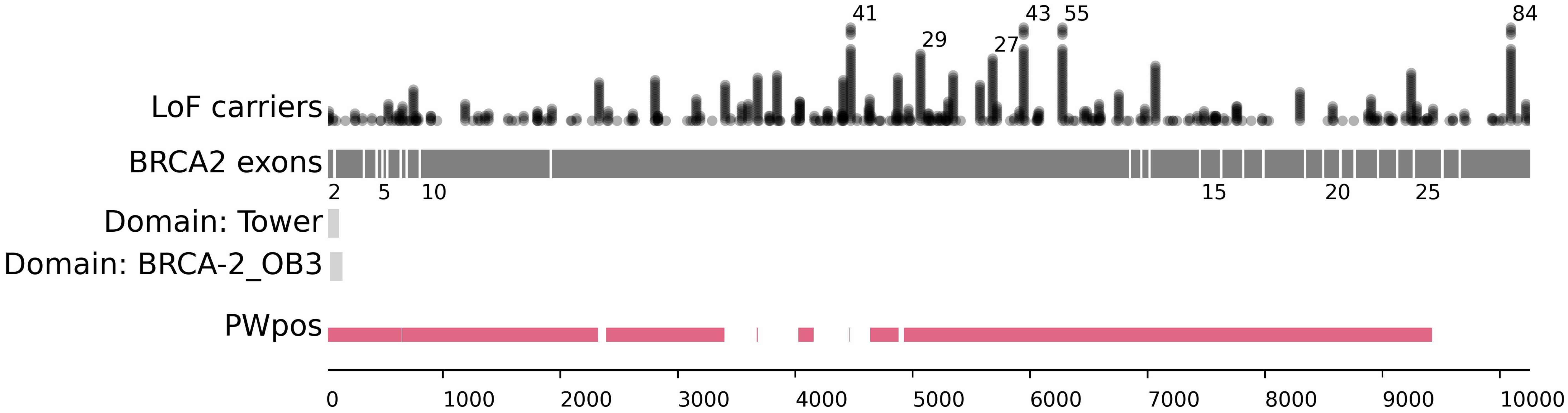

LoF carriers
41
29
27
43
55
84
BRCA2 exons
2
5
10
15
20
25
Domain: Tower
Domain: BRCA-2_OB3
PWpos
0
1000
2000
3000
4000
5000
6000
7000
8000
9000
10000

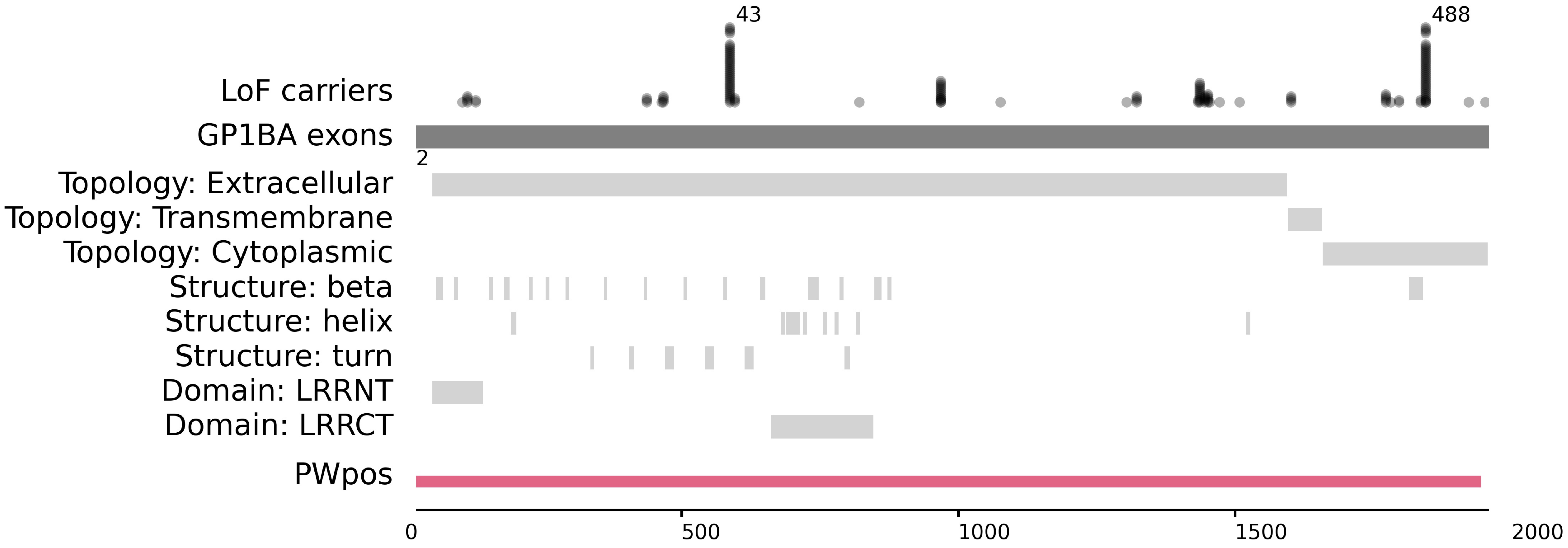

43
488
LoF carriers
GP1BA exons
2
Topology: Extracellular
Topology: Transmembrane
Topology: Cytoplasmic
Structure: beta
Structure: helix
Structure: turn
Domain: LRRNT
Domain: LRRCT
PWpos
0
500
1000
1500
2000

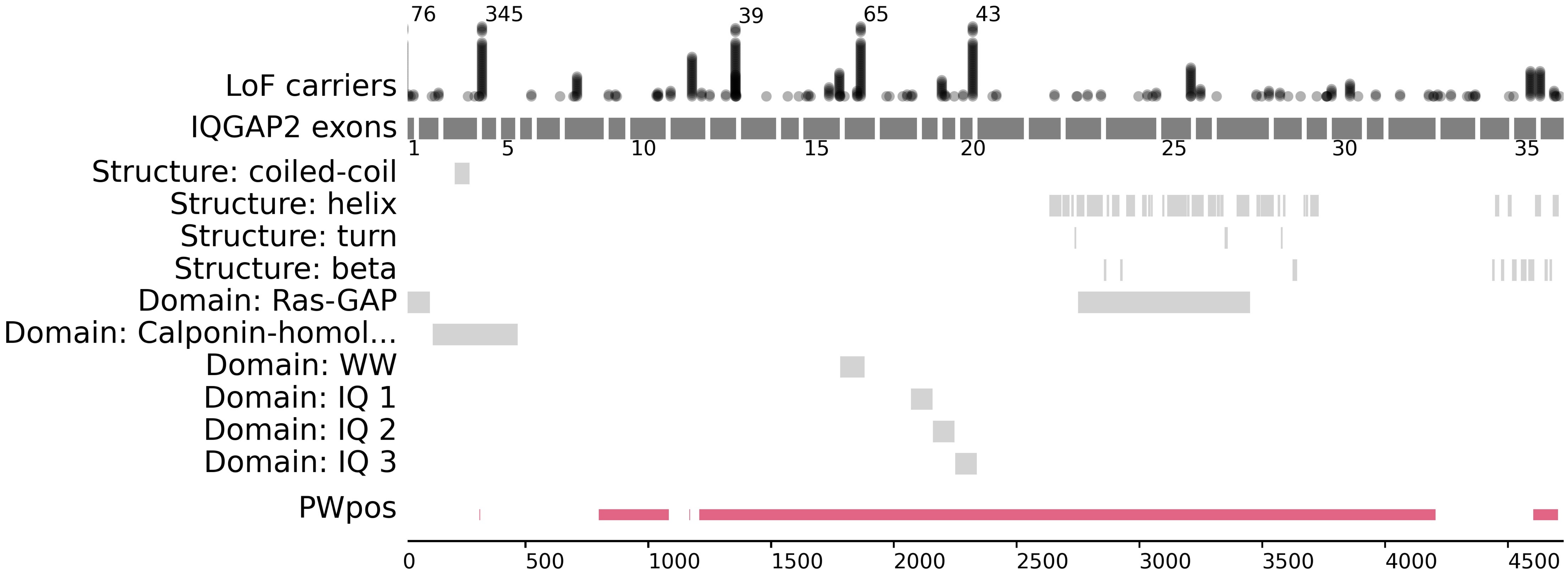
76
345
39
65
43
LoF carriers
IQGAP2 exons
1
5
10
15
20
25
30
35
Structure: coiled-coil
Structure: helix
Structure: turn
Structure: beta
Domain: Ras-GAP
Domain: Calponin-homol...
Domain: WW
Domain: IQ 1
Domain: IQ 2
Domain: IQ 3
PWpos
0
500
1000
1500
2000
2500
3000
3500
4000
4500

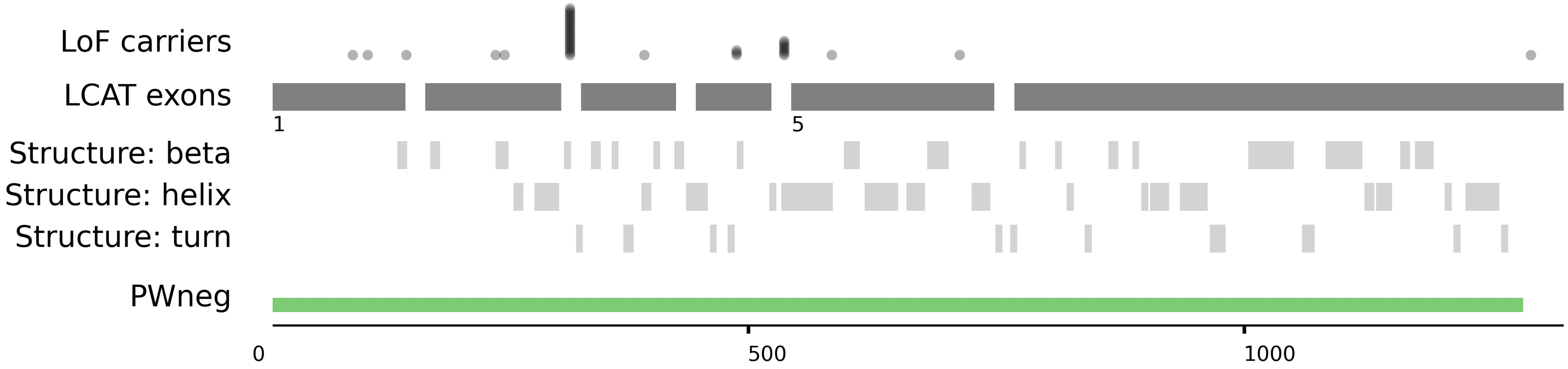

LoF carriers
LCAT exons
1
5
Structure: beta
Structure: helix
Structure: turn
PWneg
0
500
1000

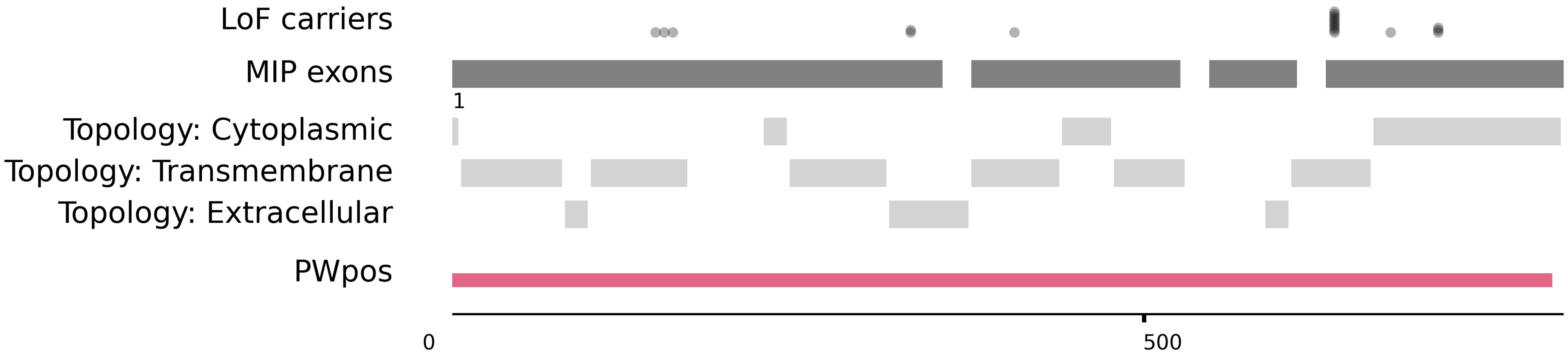

LoF carriers
MIP exons
1
Topology: Cytoplasmic
Topology: Transmembrane
Topology: Extracellular
PWpos
0
500

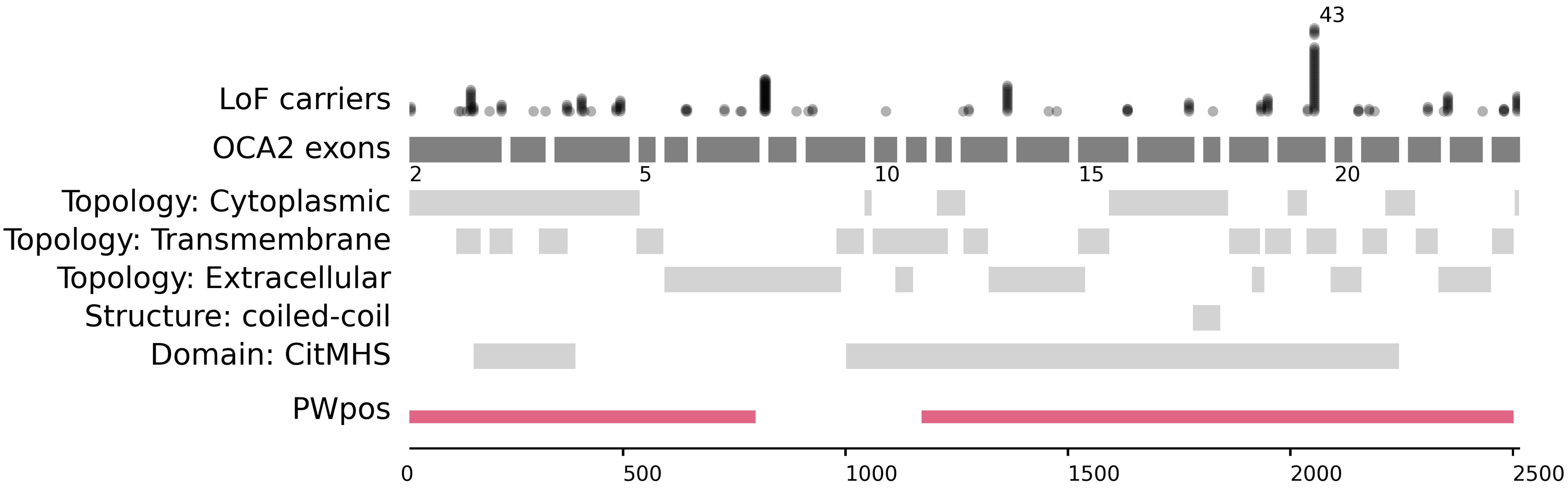

43
LoF carriers
OCA2 exons
2
5
10
15
20
Topology: Cytoplasmic
Topology: Transmembrane
Topology: Extracellular
Structure: coiled-coil
Domain: CitMHS
PWpos
0
500
1000
1500
2000
2500

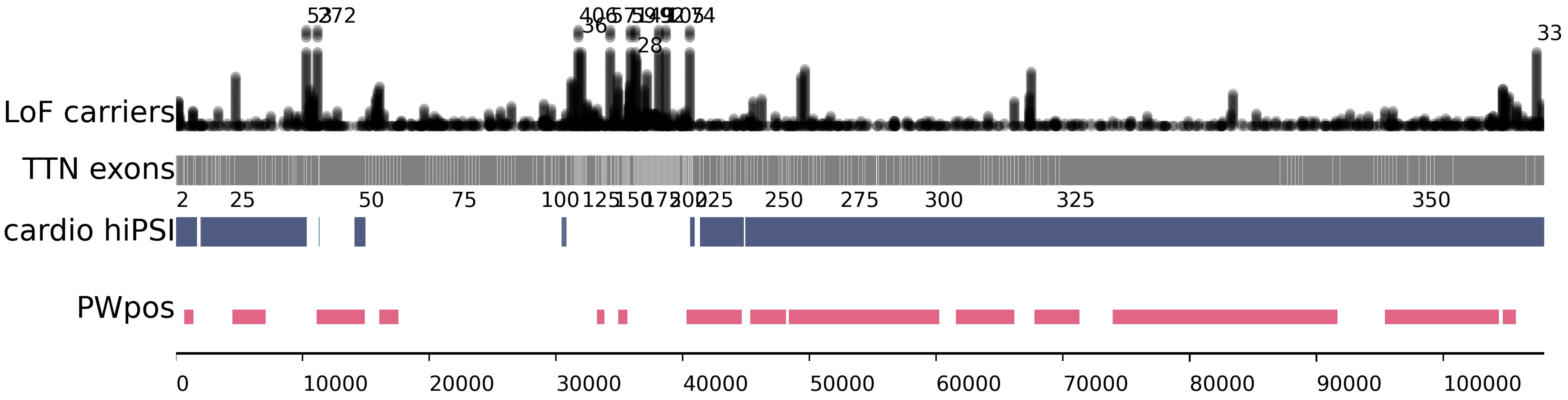

LoF carriers
TTN exons
cardio hiPSI
PWpos
2
25
50
75
100
125
150
175
200
225
250
275
300
325
350
0
10000
20000
30000
40000
50000
60000
70000
80000
90000
100000
36
28
33

**Figure S4. Power Window results for the 29 significant models.** Tracks are drawn against each gene's canonical coding transcript. Coding position is shown at the bottom scale. Carriers: each dot represents an individual carrier and are stacked for each carrier at a given position. For brevity, carriers are trimmed to 35 and total number of carriers is indicated when total carriers>40. Exons: exons (dark grey to scale; introns not to scale). Exon number indicated below exon track. Topology, secondary structures and major domains are annotated according to UniProt. PWpos: merger of all significant windows with a positive direction of effect (beta>0.5 or OR>cutoff for that gene; pink). PWneg: merger of all significant windows with a negative direction of effect (beta<-0.5; green). Significant associations are determined as indicated in the main text.

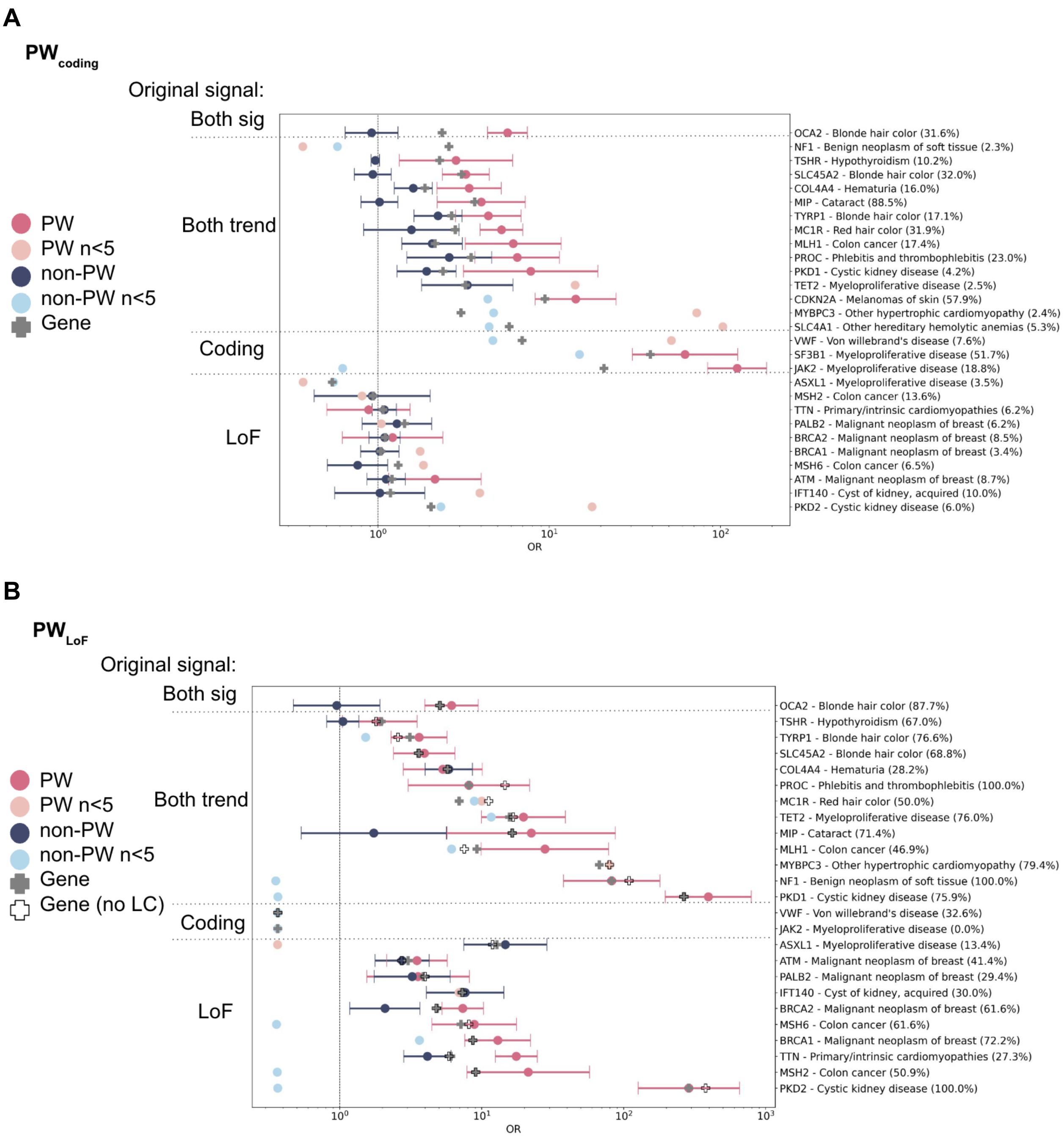


**Figure S5. Performance of Power Window for all binary traits in the 128K UKB test set**. A) Coding model B) LoF model. The odds ratios for PW models are shown in pink, and the excluded regions are shown in blue. The gene-phenotype label includes in parentheses an indication of what percent of rare variant carriers in the gene were included in the PW model. For brevity, confidence intervals are only shown when there are at least 5 case carriers: datapoints from <5 carriers are not well supported and require additional data to confirm. The genes are split into those that originally showed genome-wide significant signals for both a coding and LoF model (p<5e-8; “Both sig”), those that were genome-wide significant for either coding or LoF and showed some association with the other model (p<0.05; “Both trend”), and those that showed association with only the coding or only the LoF model at the level of the whole gene. Two whole-gene models are shown: gray filled cross, which includes all qualifying variants in the gene, and black empty cross, in which LOFTEE LC variants are excluded.

A

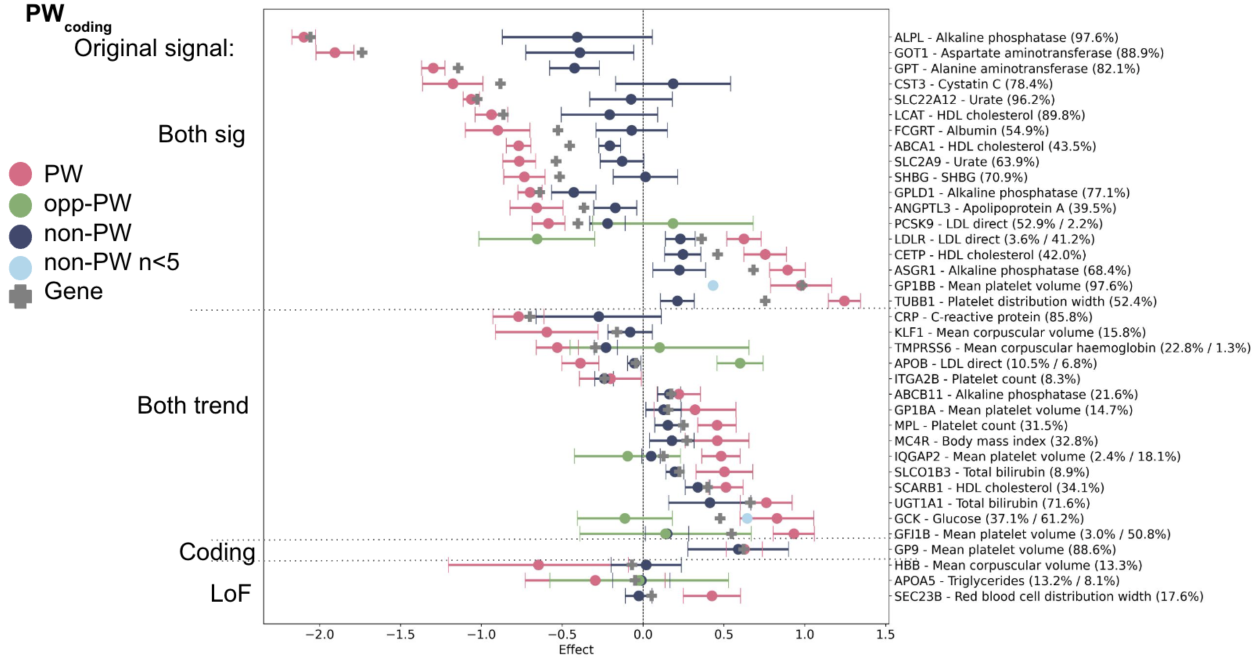


B

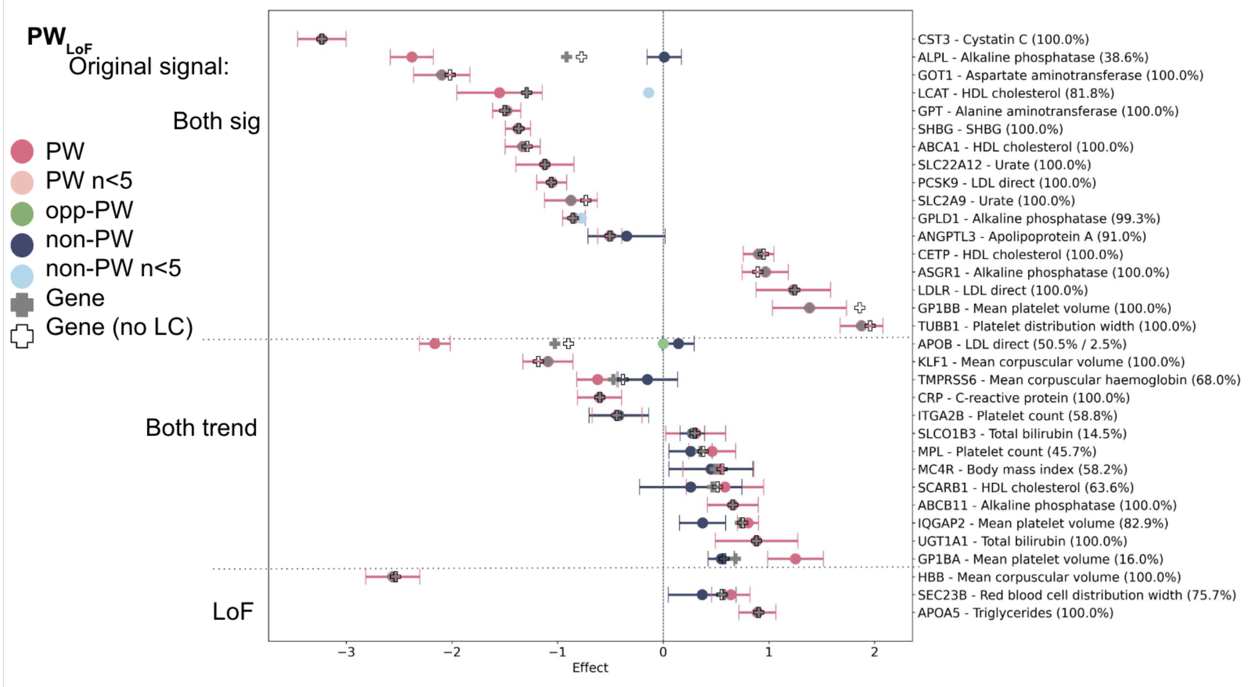


**Figure S6. Performance of Power Window for all quantitative traits in the 128K UKB test set**. A) Coding model B) LoF model. The effect sizes (normalized phenotypes) for PW models are shown in pink, and the excluded regions are shown in blue. The gene-phenotype label includes in parentheses an indication of what percent of rare variant carriers in the gene were included in the PW model (when there are 2 models for a gene, the percent shown is for negative model / positive model). When a model for the opposite direction of the main effect was built (opp-PW), it is shown in green. For brevity, confidence intervals are only shown when there are at least 5 carriers: datapoints from <5 carriers are not well supported and require additional data to confirm. The genes are split into those that originally showed genome-wide significant signals for both a coding and LoF model (p<5e-8; “Both sig”), those that were genome-wide significant for either coding or LoF and showed some association with the other model ($p<0.05$; “Both trend”), and those that showed association with only the coding or only the LoF model at the level of the whole gene. Two whole-gene models are shown: gray filled cross, which includes all qualifying variants in the gene, and black empty cross, in which LOFTEE LC variants are excluded.

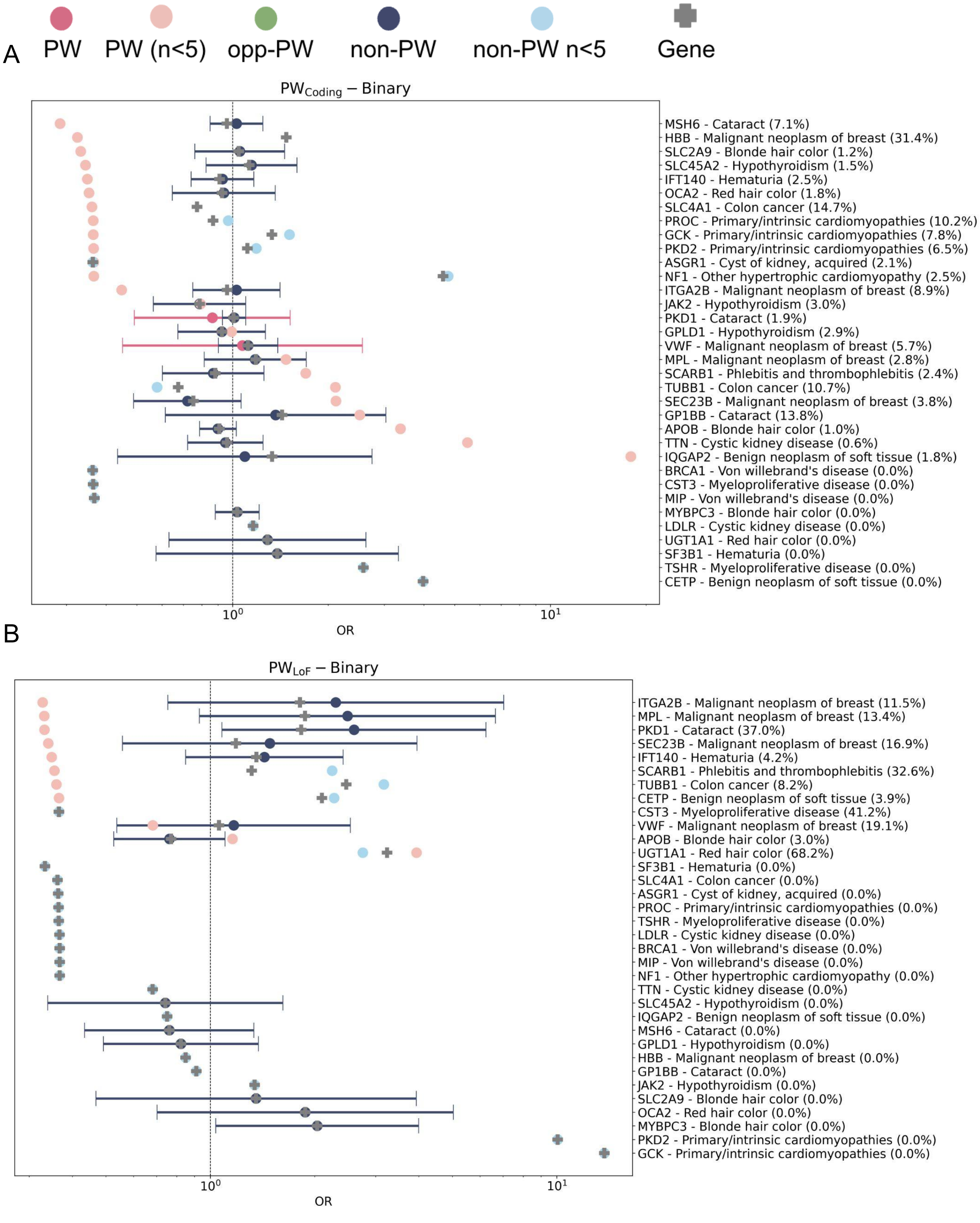

PW
PW (n<5)
opp-PW
non-PW
non-PW n<5
Gene
A
PW$_{Coding}$ – Binary
MSH6 - Cataract (7.1%)
HBB - Malignant neoplasm of breast (31.4%)
SLC2A9 - Blonde hair color (1.2%)
SLC45A2 - Hypothyroidism (1.5%)
IFT140 - Hematuria (2.5%)
OCA2 - Red hair color (1.8%)
SLC4A1 - Colon cancer (14.7%)
PROC - Primary/intrinsic cardiomyopathies (10.2%)
GCK - Primary/intrinsic cardiomyopathies (7.8%)
PKD2 - Primary/intrinsic cardiomyopathies (6.5%)
ASGR1 - Cyst of kidney, acquired (2.1%)
NF1 - Other hypertrophic cardiomyopathy (2.5%)
ITGA2B - Malignant neoplasm of breast (8.9%)
JAK2 - Hypothyroidism (3.0%)
PKD1 - Cataract (1.9%)
GPLD1 - Hypothyroidism (2.9%)
VWF - Malignant neoplasm of breast (5.7%)
MPL - Malignant neoplasm of breast (2.8%)
SCARB1 - Phlebitis and thrombophlebitis (2.4%)
TUBB1 - Colon cancer (10.7%)
SEC23B - Malignant neoplasm of breast (3.8%)
GP1BB - Cataract (13.8%)
APOB - Blonde hair color (1.0%)
TTN - Cystic kidney disease (0.6%)
IQGAP2 - Benign neoplasm of soft tissue (1.8%)
BRCA1 - Von willebrand's disease (0.0%)
CST3 - Myeloproliferative disease (0.0%)
MIP - Von willebrand's disease (0.0%)
MYBPC3 - Blonde hair color (0.0%)
LDLR - Cystic kidney disease (0.0%)
UGT1A1 - Red hair color (0.0%)
SF3B1 - Hematuria (0.0%)
TSHR - Myeloproliferative disease (0.0%)
CETP - Benign neoplasm of soft tissue (0.0%)
$10^0$
$10^1$
OR
B
PW$_{LoF}$ – Binary
ITGA2B - Malignant neoplasm of breast (11.5%)
MPL - Malignant neoplasm of breast (13.4%)
PKD1 - Cataract (37.0%)
SEC23B - Malignant neoplasm of breast (16.9%)
IFT140 - Hematuria (4.2%)
SCARB1 - Phlebitis and thrombophlebitis (32.6%)
TUBB1 - Colon cancer (8.2%)
CETP - Benign neoplasm of soft tissue (3.9%)
CST3 - Myeloproliferative disease (41.2%)
VWF - Malignant neoplasm of breast (19.1%)
APOB - Blonde hair color (3.0%)
UGT1A1 - Red hair color (68.2%)
SF3B1 - Hematuria (0.0%)
SLC4A1 - Colon cancer (0.0%)
ASGR1 - Cyst of kidney, acquired (0.0%)
PROC - Primary/intrinsic cardiomyopathies (0.0%)
TSHR - Myeloproliferative disease (0.0%)
LDLR - Cystic kidney disease (0.0%)
BRCA1 - Von willebrand's disease (0.0%)
MIP - Von willebrand's disease (0.0%)
NF1 - Other hypertrophic cardiomyopathy (0.0%)
TTN - Cystic kidney disease (0.0%)
SLC45A2 - Hypothyroidism (0.0%)
IQGAP2 - Benign neoplasm of soft tissue (0.0%)
MSH6 - Cataract (0.0%)
GPLD1 - Hypothyroidism (0.0%)
HBB - Malignant neoplasm of breast (0.0%)
GP1BB - Cataract (0.0%)
JAK2 - Hypothyroidism (0.0%)
SLC2A9 - Blonde hair color (0.0%)
OCA2 - Red hair color (0.0%)
MYBPC3 - Blonde hair color (0.0%)
PKD2 - Primary/intrinsic cardiomyopathies (0.0%)
GCK - Primary/intrinsic cardiomyopathies (0.0%)
$10^0$
$10^1$
OR

C

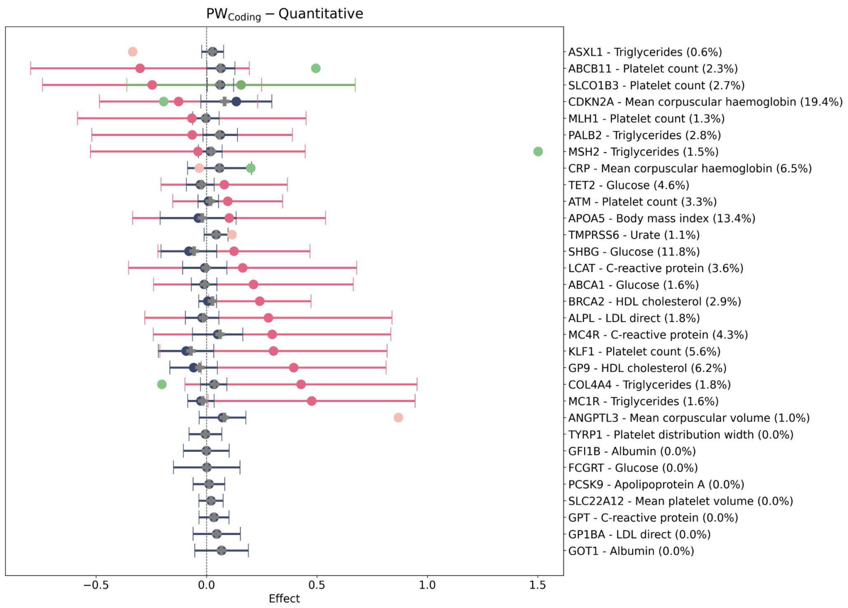


D

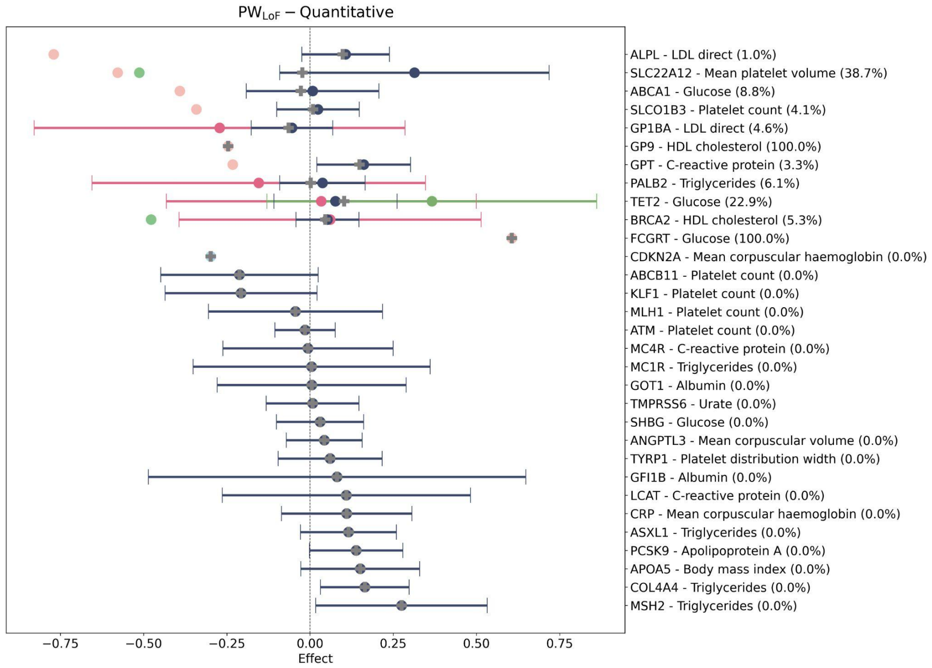


**Figure S7. Performance of Power Window for phenotype negative control in the 128K UKB test set**. Here, the same phenotypes and genes from the main analysis were randomly reassigned, so that the gene was no longer expected to show an association with the trait. Models were built for the negative control traits in the 300k UKB cohort and then examined in the 128k followup samples. The number in parentheses indicates the percentage of variant carriers in that gene that were kept in the PW model. For many combinations, no models were able to be built (rows without pink dots or lines), and the models that were built did not predict the phenotypes in the test set.

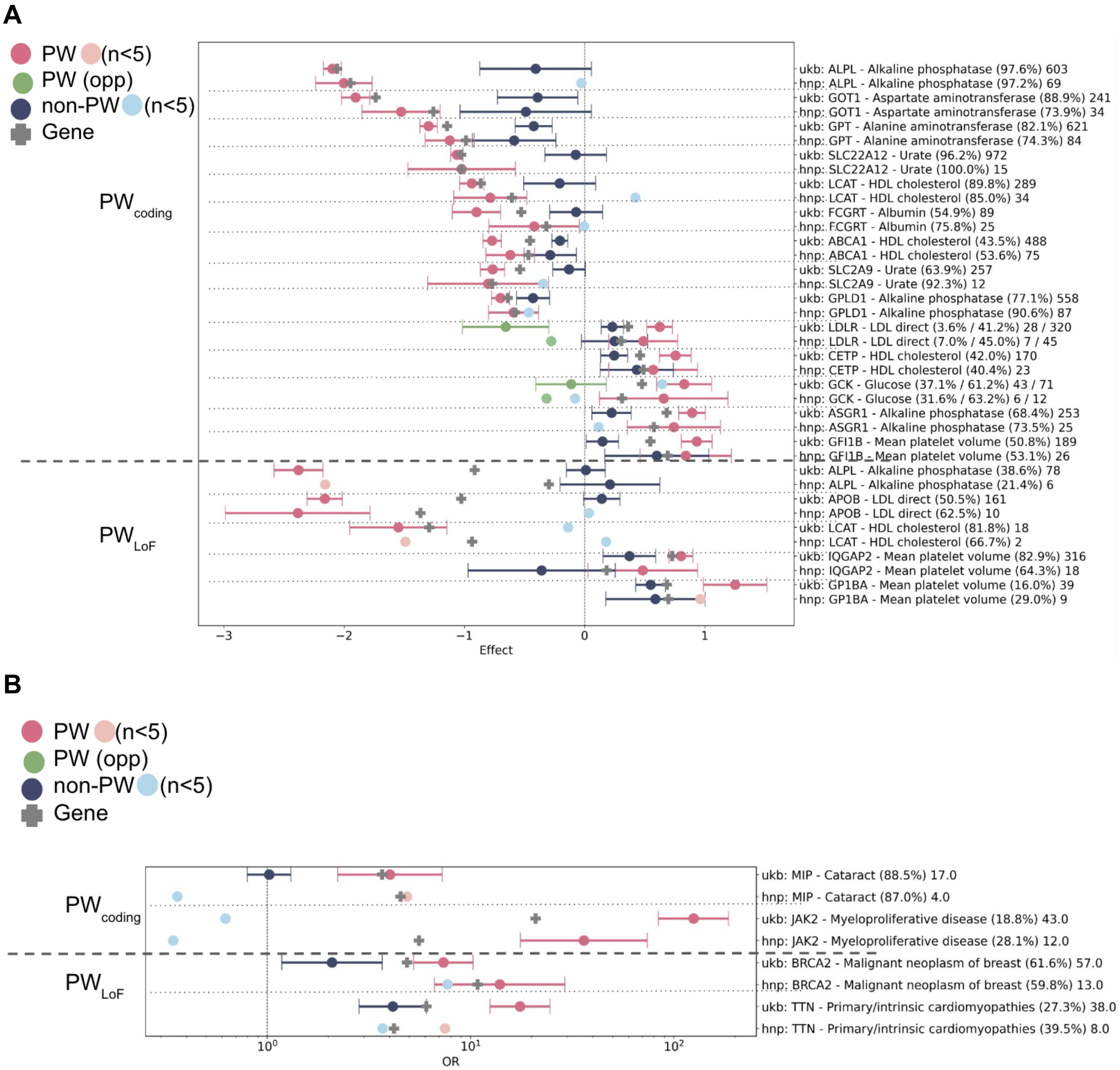


**Figure S8 Replication in the HNP cohort.** A) Quantitative traits. The effect sizes (normalized phenotypes) for PW models are shown in pink, and the excluded regions (non-PW) are shown in blue. When a model for the opposite direction of the main effect was built (PW-opp), it is shown in green. B) Binary traits. The odds ratios for PW models are shown in pink, and the excluded regions (non-PW) are shown in blue. The gene-phenotype label includes in parentheses an indication of what percent of rare variant carriers in the gene were included in the PW model (when there are 2 models for a gene, the percent shown is for negative model / positive model). For brevity, confidence intervals are only shown when there are at least 5 carriers (5 case carriers for binary traits): datapoints from <5 carriers are not well supported and require additional data to confirm. Only models that were significant in the UKB128k test cohort are tested, and at least 5 case carriers expected in the HNP cohort (UKB case carrier frequency * HNP n cases) is required for binary traits. The UKB data shown are from the 128k test cohort. The total number of PW variant carriers is shown at the end of each row.

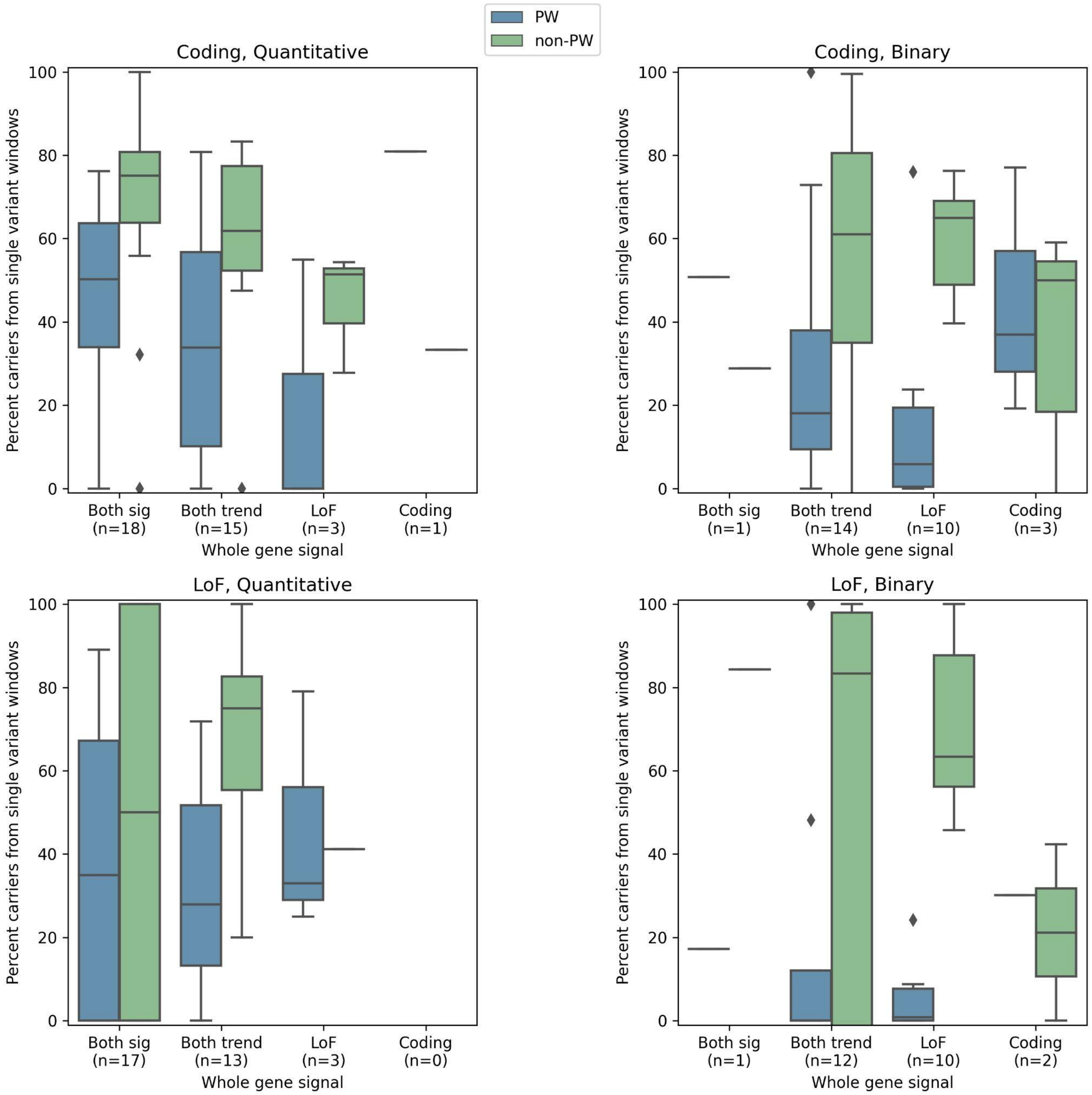


**Figure S9. Percent carriers in PW and non-PW models that are from single variant windows.** Variants with a frequency above the cutoff (20 carriers) were pulled out into their own analyses, separate from the sliding windows, and were called single variant windows. For PW models, single variant windows made up a mean of 34% / 28% (range 0-100%) of the carriers in the $PW_{Coding}$ / $PW_{LoF}$ models, compared to 59% / 56% (range 0-100%) of the non-PW carriers.

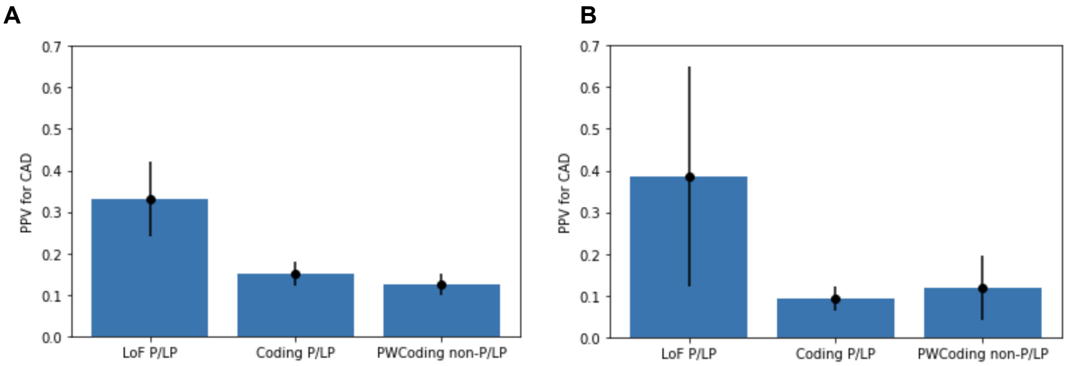


**Figure S10. Penetrance of LDLR variants for CAD (coronary artery disease) for A) UKB and B) HNP.** LoF variants have a PPV of 30-40% in these cohorts, while Coding variants had PPVs of 10-15% when they were known P/LP variants from ClinVar or were novel $PW_{coding}$ variants. The PPV was 6-8% in rare coding variants that were non-P/LP and non-$PW_{Coding}$.
